# HIV seroprevalence, incidence, and viral suppression among Ugandan female bar workers: a population-based study

**DOI:** 10.1101/2025.03.22.25324404

**Authors:** Xinyi Feng, Gertrude Nakigozi, Eshan U. Patel, Caitlin E. Kennedy, Slisha Shrestha, Fred Nalugoda, Godfrey Kigozi, Robert Ssekubugu, Larry W Chang, Andrea L. Wirtz, Hadijja Nakawooya, Grace Kigozi, Ronald M Galiwango, Steven J Reynolds, Joseph Kagaayi, Aaron A. R. Tobian, M. Kate Grabowski

**Affiliations:** Department of Pathology, Johns Hopkins University School of Medicine, Baltimore, MD, USA; Rakai Health Sciences Program, Kalisizo, Uganda; Department of Epidemiology, Johns Hopkins School of Public Health, Baltimore, MD, USA; Department of International Health, Johns Hopkins Bloomberg School of Public Health, Baltimore, MD, USA; Division of Infectious Diseases, Department of Medicine, Johns Hopkins School of Medicine, Baltimore, MD, USA; National Institute of Allergy and Infectious Diseases, Bethesda, Maryland, USA

## Abstract

**Background:** Prior studies have linked female bar and sex work in Africa. However, population-level data on HIV burden among female bar workers (FBWs) in African settings are rare.

**Methods:** We used five survey rounds of data collected between 2011 and 2020 from the Rakai Community Cohort Study, a population-based HIV surveillance cohort in 36 inland agrarian and trading communities (HIV prevalence ~12%) and four Lake Victoria fishing communities (~36%) in southern Uganda. Women reporting bar work as a primary or secondary occupation were identified and compared to non-FBWs. Primary outcomes included laboratory-confirmed HIV seropositivity, incident infection, viral suppression (<200 copies/ml) among women with HIV, and population prevalence of viremia. Prevalence ratios (PRs) and incidence rate ratios (IRRs) were estimated using Poisson regression models with 95% confidence intervals (95%CI).

**Findings:** A total of 23,556 female participants contributed 52,708 person visits. Overall, 1,205 (5.1%) women self-identified as FBWs. FBWs had significantly higher baseline HIV seroprevalence compared to non-FBWs (51.9% vs. 18.5%,PR=2.81, 95%CI=2.64-2.95). 356 HIV incident events occurred over 39,228 years of participant follow-up. Incidence among FBWs was 2.49/100 person-years versus 0.87/100 person-years among non-FBWs (age-adjusted IRR=3.64,95%CI=2.33-5.42). While HIV viral suppression was similar among participants living with HIV regardless of FBW status, the population prevalence of HIV viremia among FBWs was 1.69 times higher compared to non-FBWs, adjusting for age and community type (95%CI=1.38-2.08). Among 179 HIV seronegative FBWs surveyed in 2018-20, 79.9% (143/179) were aware of PrEP, while only 13.4% (24/179) had ever used it, and just 2.8% (5/179) were current users.

**Interpretation:** FBWs in Uganda experience substantially higher HIV burden and acquisition risk compared to the general population. Tailored prevention strategies like prioritizing their HIV service delivery may reduce HIV incidence among FBWs and their partners.

**Funding:** National Institute of Allergy and Infectious Diseases, National Institutes of Health

## Introduction

Despite Eastern and Southern Africa (ESA) having the largest reduction in new HIV cases globally over the past decade, HIV incidence remains highest in ESA.^1^ As incidence declines, new infections are expected to become increasingly concentrated in populations disproportionately affected by HIV, including female sex workers (FSWs) and their sexual partners. Mathematical models estimate that these groups account for up to 25% of recent infections, with an incidence rate ~5 times greater than the general population in ESA.^2^ Consequently, FSWs are recognized as a key population and have been prioritized for HIV services, including pre-exposure prophylaxis (PrEP), but stigma and legal barriers surrounding sex work hinder research and the effective delivery of interventions.^3^

Bar work, which involves employment in establishments serving alcoholic beverages, is a common source of employment for women in ESA.^4^ Many female bar workers (FBWs) also engage in sex work to supplement their income.^5,6^ Although FBWs do not typically self-identify as FSWs, they are considered an informal subgroup of FSWs in some ESA contexts.^5,7^ Therefore, HIV surveillance among FBWs may offer important insights into HIV dynamics among FSWs, especially among those less likely to self-disclose sex work. Beyond sex work, bar work carries independent risks for HIV, including increased mobility, alcohol and drug use, stigmatization, and exposure to violence.^5,8–11^

Unlike FSWs, FBWs are not formally recognized as key or priority populations in HIV epidemic control programs. This is partly because data on HIV burden among FBWs are rare and limited to cross-sectional studies.^4^ Only five studies have measured HIV seroprevalence among African FBWs, with estimates ranging from 7.1% to 68%; only study one in the last 20 years.^7,12–15^ This most recent study from Tanzania, had the lowest seroprevalence estimate and included just 66 FBWs.^15^ None of these studies systematically compared HIV seroprevalence among FBWs with that of women not involved in bar work in the same survey, nor did they assess HIV incidence or viral suppression. Such comparisons are crucial for estimating the HIV burden faced by FBWs and developing appropriately tailored HIV services.

In this study, we used longitudinal data from Rakai, Uganda to assess HIV seroprevalence, incidence, viral suppression, and population prevalence of viremia among FBWs and compared it with that in the local female population not engaged in bar work (i.e., non-FBWs). Additionally, we used longitudinal data to examine patterns of entry into and exit bar work over time, and evaluated the associations between bar work exposure prior to and after entry with HIV serostatus. These data may address critical gaps in HIV surveillance among FBWs in ESA and provide important insights into the relationship between bar work and HIV burden, which may help guide future targeted outreach and interventions for persons most vulnerable to infection in the context of a declining HIV epidemic.

## Methods

### Study design and population

The Rakai Community Cohort Study (RCCS), conducted by the Rakai Health Sciences Program (RHSP), is an open population-based longitudinal HIV surveillance study in southern Uganda.^16^ The RCCS includes both a household census and survey in four Lake Victoria fishing communities with high adult HIV seroprevalence (~40%) and 36 inland agrarian and trading communities with relatively moderate seroprevalence (~12%).^17^ The census documents all individuals residing in households within community surveillance boundaries irrespective of age or residence status, as well as migration events (in- and out-migration), births, and deaths. The survey is conducted ~2 weeks after the census and includes community residents (>1 month with intention to stay) aged 15-49 years with the capacity to provide informed consent. Participants’ socioeconomic status (SES) is assessed by a household asset-based measure and categorized into quartiles after standardization by Z-scores.^18^ Participants are provided with HIV testing and counseling services, and persons living with HIV are referred for HIV treatment services.

Here, we conducted a secondary data analysis of RCCS data obtained from female participants aged 15-49 between Aug 10, 2011 and November 06, 2020. The analysis period included five survey rounds, herein denoted RCCS survey rounds 15 (August 10, 2011–July 5, 2013), 16 (July 8, 2013–January 30, 2015), 17 (February 23, 2015–September 2, 2016), 18 (Oct 3, 2016–May 22, 2018), and 19 (June 19, 2018–November 6, 2020).

### Measurement and classification of female bar worker status

RCCS data on self-reported primary and secondary occupations were used to determine FBW status and the primary exposure of interest. During the interview, participants were asked the question: *“What kind of work do you do, or what kind of activities keep you busy during an average day, whether you get money for them or not?”* We defined female participants who reported being either a bar worker or owners as either their primary or secondary occupation as FBW at each study visit. Participants who did not report bar work at a visit were defined as non-FBWs. We considered FBW status as both a time-varying (i.e., FBW versus non-FBW during a survey round) and invariant (i.e., never-versus ever-FBW during the study period) exposure. We assessed baseline HIV seroprevalence by participants’ ever-FBW status. FBW status at the current visit was employed as the main exposure for estimates of viral suppression and population prevalence of HIV viremia. Additionally, the recent FBW status, defined as self-reported bar work at either the start or end of a visit interval, was used to estimate the incidence of HIV infection. There were 2,609 participants not surveyed about their occupation in survey round 16 due to protocol changes in the questionnaire during that survey round. For these women, we imputed their occupation using the most recent reported occupation at either subsequent or prior visits. There were 116 participants who only took part in survey round 16, and these women were considered non-FBWs (details in **Supplementary_Statistical_Methods**).

### Measurement of HIV-related outcomes

Primary study outcomes included HIV seroprevalence, HIV seroconversion (i.e., incident infection), HIV viral suppression, and HIV viremia. HIV serostatus was assessed from participants’ venous blood samples using a validated parallel three-test rapid HIV testing algorithm (Determine, Stat-Pak, and Uni-Gold) and confirmed by laboratory tests via enzyme immunoassays (Murex HIV-1, 2.O).^19^ HIV viral load was determined using stored serum from participants with HIV using the Abbott RealTime HIV-1 Assay including m2000sp (sample preparation instrument) and the m2000rt (RealTime PCR analyzer) (Abbott Molecular, Inc., Des Plaines, IL).^20^ HIV seroconversion events were defined as instances in which persons tested HIV seropositive for the first time following an HIV-seronegative result at the prior study visit (allowing for up to one missed visit). HIV viral suppression was defined as having an HIV viral load <200 copies/mL. Conversely, HIV viremia was defined as having an HIV viral load ≥200 copies/mL.

### Statistical Analysis

We assessed baseline sociodemographic and sexual behavior characteristics by self-reported FBW status at any visit using descriptive statistics and Pearson’s chi-squared tests. For never-FBWs, we defined their baseline visit as the first study visit during the study period, whereas for ever-FBWs, we used the first visit at which they self-reported bar work.

Baseline five-year age-specific HIV seroprevalence among never-FBWs and ever-FBWs was evaluated within inland and fishing communities separately. The Clopper-Pearson method was used to compute corresponding 95% confidence intervals (95%CIs).^21^ We assessed HIV seroprevalence by whether or not bar work was reported as either a primary or secondary occupation or both. For FBWs who did not self-report bar work at their first visit (i.e., future FBWs), we assessed HIV seroprevalence at that visit and compared it to the HIV seroprevalence at the first visits of never-FBWs and FBWs who reported bar work at their first visit. (i.e., current FBWs). We also assessed HIV seroprevalence at the final visit among FBWs who no longer reported bar work (i.e., previous FBWs) and compared it to the HIV seroprevalence at the final visits of never-FBWs and FBWs who reported bar work at their last visit. Additionally, we measured HIV seroprevalence among FBWs at their final visit by the total number of visits they reported being an FBW during the study period.

Subsequently, we evaluated sociodemographic and behavioral characteristics associated with baseline HIV seroprevalence separately for ever-FBW and never-FBW using modified Poisson regression with robust standard errors, with associations reported as prevalence ratios (PRs) with 95%CIs in univariable and multivariable regression analyses. Multivariate models included adjustment for age (model 2), age and other demographics (model 3), and age, demographics, and sexual behaviors (model 4).

The unit of analysis for estimating HIV incidence was person-years of follow-up between surveys among individuals who initially tested negative for HIV and provided data for at least two consecutive visits (allowing for up to one missed visit). In our primary analyses of HIV incidence, we examined the relationship between recent bar work status and HIV seroconversion, treating bar work as a time-varying exposure (recent FBWs), as we hypothesized that HIV acquisition may be influenced by current bar work status. For sensitivity analyses, we considered bar work as a time-invariant exposure (ever-FBW), to assess whether having ever engaged in bar work is associated with a higher risk of HIV acquisition, reflecting the possibility that this population may carry an elevated risk of HIV irrespective of current bar work involvement. Seroconversion events were assumed to occur at the midpoint of the interval between visits. Incidence rates (IRs) and corresponding 95%CIs were presented as the number of new HIV seroconversions per 100 person-years. Poisson regression models were utilized to estimate incidence rate ratios (IRRs) of HIV seroconversion comparing FBWs to non-FBWs. Both unadjusted and age-adjusted IRRs were computed, and stratified analyses were performed by community type and calendar time (survey rounds 15-17 and 18-19). To address potential selection biases resulting from differential loss to follow-up, we also calculated IRRs incorporating stabilized inverse-probability-of-censoring weighting (details in **Supplementary_Statistical_Methods**).

Among women with HIV, we assessed viral suppression by FBW status at each survey round, starting from round 16 (viral load data were unavailable for round 15). We compared viral suppression levels among FBWs and non-FBWs across all surveys using Poisson regression models with generalized estimating equations (GEE) and an exchangeable correlation structure. These analyses were further adjusted for age and stratified by community type. We also calculated the population prevalence of HIV viremia irrespective of HIV serostatus by survey round and FBW status. Within each survey round, population prevalence ratios of HIV viremia comparing FBWs to non-FBWs were then estimated using multivariable Poisson regression models with robust standard errors, adjusting for participants’ age and community type. Additionally, we estimated the overall population prevalence ratio of HIV viremia, comparing FBWs to non-FBWs across all surveys using Poisson regression with GEE and an exchangeable correlation structure.

Lastly, we assessed awareness of HIV serostatus among women with HIV and PrEP awareness and use (current and ever) among HIV-seronegative women at the final survey round (round 19), during which these variables were measured. Data were summarized by frequencies and percentages, with Pearson’s chi-squared tests to assess statistically significant differences by FBW status.

Statistical analyses were carried out using Stata 15.1 (Stata Corp, College Station, TX) and R 4.2.1 (R statistical software, 2022). All p-values were two-sided; we considered a *p*-value of <0.05 statistically significant.

### Ethics

This activity was reviewed and approved by Uganda Virus Research Institute, Research and Ethics Committee (approval number GC/127/19/05/654), and the Johns Hopkins School of Medicine Institutional Review Board. All participants provided written informed consent.

### Role of the funding source

The findings and conclusions in this report are those of the author(s) and do not necessarily represent the official position of the funding agencies.

## Results

### Study population characteristics

Of the 36,268 eligible females in the five RCCS survey rounds, 23,556 (64.9%) females participated and contributed 52,708 person visits (**Supplementary_Table_1, Supplementary_Figure_1**). Among them, 1,205 women (5.1%) self-identified as FBWs during one or more study visits. At baseline, ever-FBWs were typically older, less educated, lower SES, more likely to reside in fishing communities, more likely to be recent migrants, more likely to be previously married, and reported more HIV-associated sexual behaviors (e.g., transactional sex, past year and lifetime sexual partners, alcohol use before sex) compared to never-FBWs (**Table_1**).

**Table 1.** Baseline characteristics by self-reported history of bar work as a primary or secondary occupation among female participants in the Rakai Community Cohort Study, Uganda, 2011-2020 (N=23 556)

| Characteristics | Ever female bar workers<br>N=1205 | Never female bar workers<br>N=22 351 | *P*-value |
| --- | --- | --- | --- |
| **Age group** |  |  | <0.001 |
| 15-19 | 60 (5.0%) | 6460 (28.9%) |  |
| 20-24 | 168 (13.9%) | 5040 (22.5%) |  |
| 25-29 | 270 (22.4%) | 3953 (17.7%) |  |
| 30-34 | 292 (24.2%) | 2972 (13.3%) |  |
| 35-39 | 246 (20.4%) | 2041 (9.1%) |  |
| 40-44 | 117 (9.7%) | 1192 (5.3%) |  |
| 45-49 | 52 (4.3%) | 693 (3.1%) |  |
| **Community type** |  |  | <0.001 |
| Inland | 661 (54.9%) | 18 099 (81.0%) |  |
| Fishing | 544 (45.1%) | 4252 (19.0%) | <0.001 |
| **Migration status** |  |  |  |
| Long-term residents | 1018 (84.5%) | 20 402 (91.3%) | <0.001 |
| In-migrants | 187 (15.5%) | 1948 (8.7%) |  |
| Missing | 0 (0.0%) | 1 (<0.1%) |  |
| **Education attainment** |  |  | <0.001 |
| None | 144 (12.0%) | 989 (4.4%) |  |
| Primary | 835 (69.3%) | 12 195 (54.6%) |  |
| Secondary | 196 (16.3%) | 7213 (32.3%) |  |
| Technical/university | 30 (2.5%) | 1953 (8.7%) |  |
| Missing | 0 (0.0%) | 2 (<0.1%) |  |
| **Socioeconomic status** |  |  | <0.001 |
| Lowest | 251 (20.8%) | 3229 (14.4%) |  |
| Low-middle | 187 (15.5%) | 3211 (14.4%) |  |
| High-middle | 422 (35.0%) | 7334 (32.8%) |  |
| Highest | 342 (28.4%) | 8555 (38.3%) |  |
| Missing | 3 (0.2%) | 22 (0.1%) |  |
| **Marital status** |  |  | <0.001 |
| Currently married | 492 (40.8%) | 11 968 (53.5%) |  |
| Previously married | 596 (49.5%) | 3862 (17.3%) |  |
| Never married | 117 (9.7%) | 6513 (29.1%) |  |
| Missing | 0 (0.0%) | 8 (<0.1%) |  |
| **Number of lifetime sexual partners** |  |  | <0.001 |
| 0 | 4 (0.3%) | 3531 (15.8%) |  |
| 1 | 25 (2.1%) | 3720 (16.7%) |  |
| 2 | 132 (11.0%) | 5198 (23.3%) |  |
| ≥3 | 1039 (86.2%) | 9881 (44.2%) |  |
| Missing | 5 (0.4%) | 21 (0.1%) |  |
| **Number of sexual partners in the past year** |  |  | <0.001 |
| 0 | 24 (2.0%) | 3940 (17.6%) |  |
| 1 | 779 (64.6%) | 15 904 (71.2%) |  |
| 2 | 254 (21.1%) | 1947 (8.7%) |  |
| ≥3 | 148 (12.3%) | 523 (2.3%) |  |
| Missing | 0 (0.0%) | 37 (0.2%) |  |
| **Genital ulcers in the past year\*** |  |  | <0.001 |
| No | 1003 (83.2%) | 19 567 (87.5%) |  |
| Yes | 201 (16.7%) | 2781 (12.4%) |  |
| Missing | 1 (0.1%) | 3 (<0.1%) |  |
| **Transactional sex in the past year^** |  |  | <0.001 |
| No | 952 (80.6%) | 16 505 (89.8%) |  |
| Yes | 229 (19.4%) | 1869 (10.2%) |  |
| **Non-marital partnership in the past year^** |  |  | <0.001 |
| No | 424 (35.9%) | 11 366 (61.9%) |  |
| Yes | 757 (64.1%) | 7008 (38.1%) |  |
| **Consistent condom usage with nonmarital partners in the past year<sup>#</sup>** |  |  | 0.030 |
| No | 614 (81.1%) | 5445 (77.7%) |  |
| Yes | 143 (18.9%) | 1563 (22.3%) |  |
| **Alcohol use before sex in the past year<sup>&</sup>** |  |  | <0.001 |
| No | 387 (39.1%) | 12 794 (79.3%) |  |
| Yes | 604 (60.9%) | 3344 (20.7%) |  |
| **Partners' alcohol use before sex in the past year<sup>&</sup>** |  |  | <0.001 |
| No | 228 (23.0%) | 9456 (58.6%) |  |
| Yes | 763 (77.0%) | 6682 (41.4%) |  |

\* Genital ulcers in the past year for round 15-18, and genital ulcers in the past month for round 19
<sup>^</sup>Among sexually active participants in the past year (N=19 555)
<sup>#</sup>Among sexually active participants with nonmarital partners in the past year (N=7765)
<sup>&</sup>Among survey round 15-18 sexually active participants in the past year (N=17 129)

### Population prevalence of HIV

At baseline, HIV seroprevalence was significantly higher among FBWs (51.9%[625/1,205]) as compared to non-FBWs (18.5% [4,130/22,351]) (age-adjusted PR=2.17,95%CI=2.03-2.31) (**Supplementary_Table_2**). These differences were observed regardless of age or community of residence (inland communities: 45.4% vs. 13.7%, PR=3.32,95%CI=3.03-3.64; fishing communities: 59.7% vs. 39.0%, PR=1.53,95%CI=1.42-1.66) or other behavioral or demographic characteristics (**Figure_1, Supplementary_Table_3**). Moreover, differences in seroprevalence by FBW status persisted through the most recent survey period (**Supplementary_Figure_2, Supplementary_Figure_3**).

**Figure 1.**
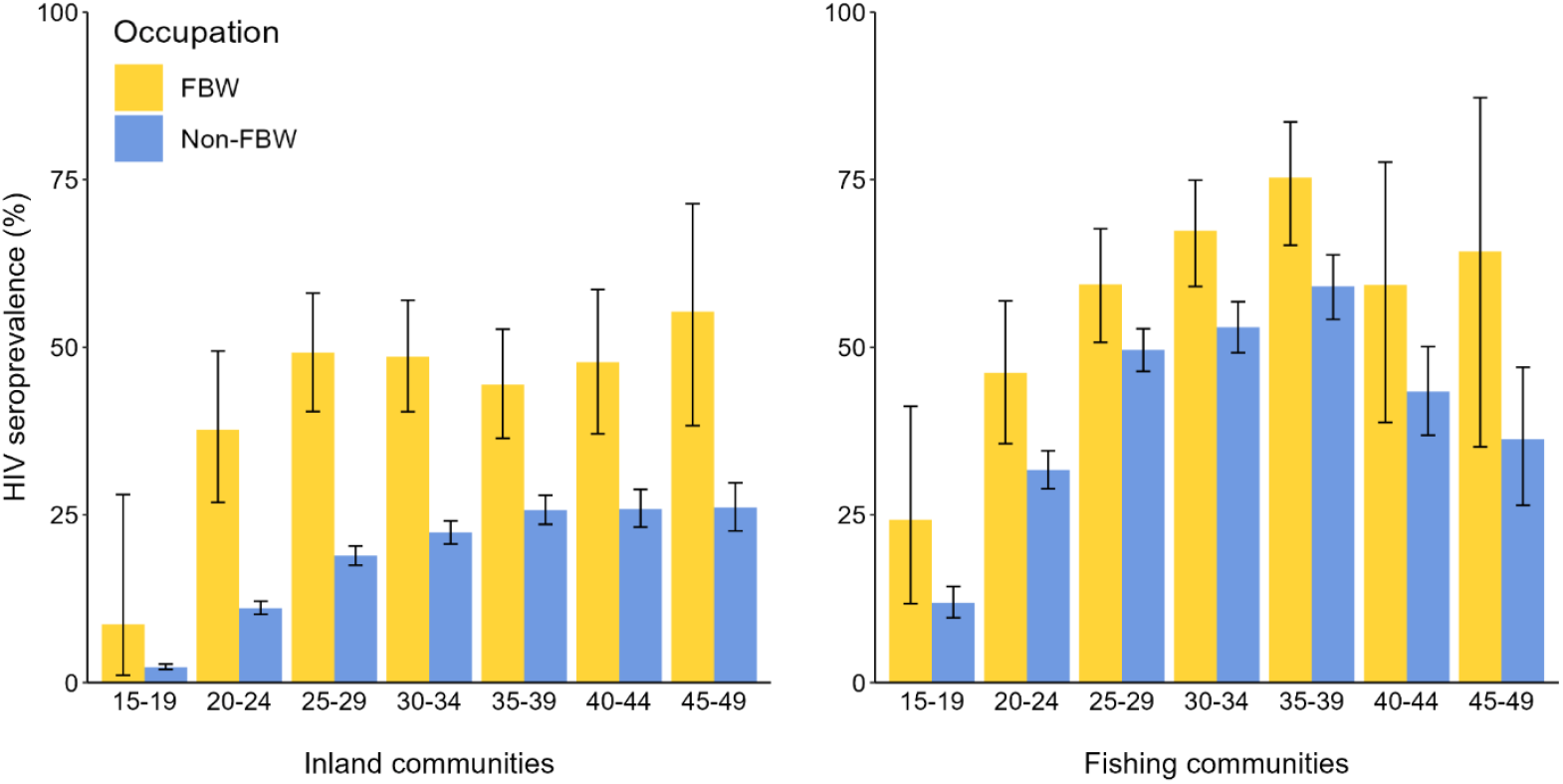
Age-specific baseline HIV seroprevalence by self-reported history of bar work as a primary or secondary occupation in the inland and fishing communities among female participants of Rakai Community Cohort Study, Uganda, 2011-2020 (N=23 556) Abbreviations: FBW, female bar worker; HIV, human immunodeficiency virus

**Table 2** shows sociodemographic and behavioral factors associated with seroprevalent HIV infection at baseline stratified by FBW status. Factors associated with HIV seroprevalence were similar for FBWs and non-FBWs. However, associations between these factors and HIV seroprevalence were typically stronger among non-FBW.

**Table 2.**
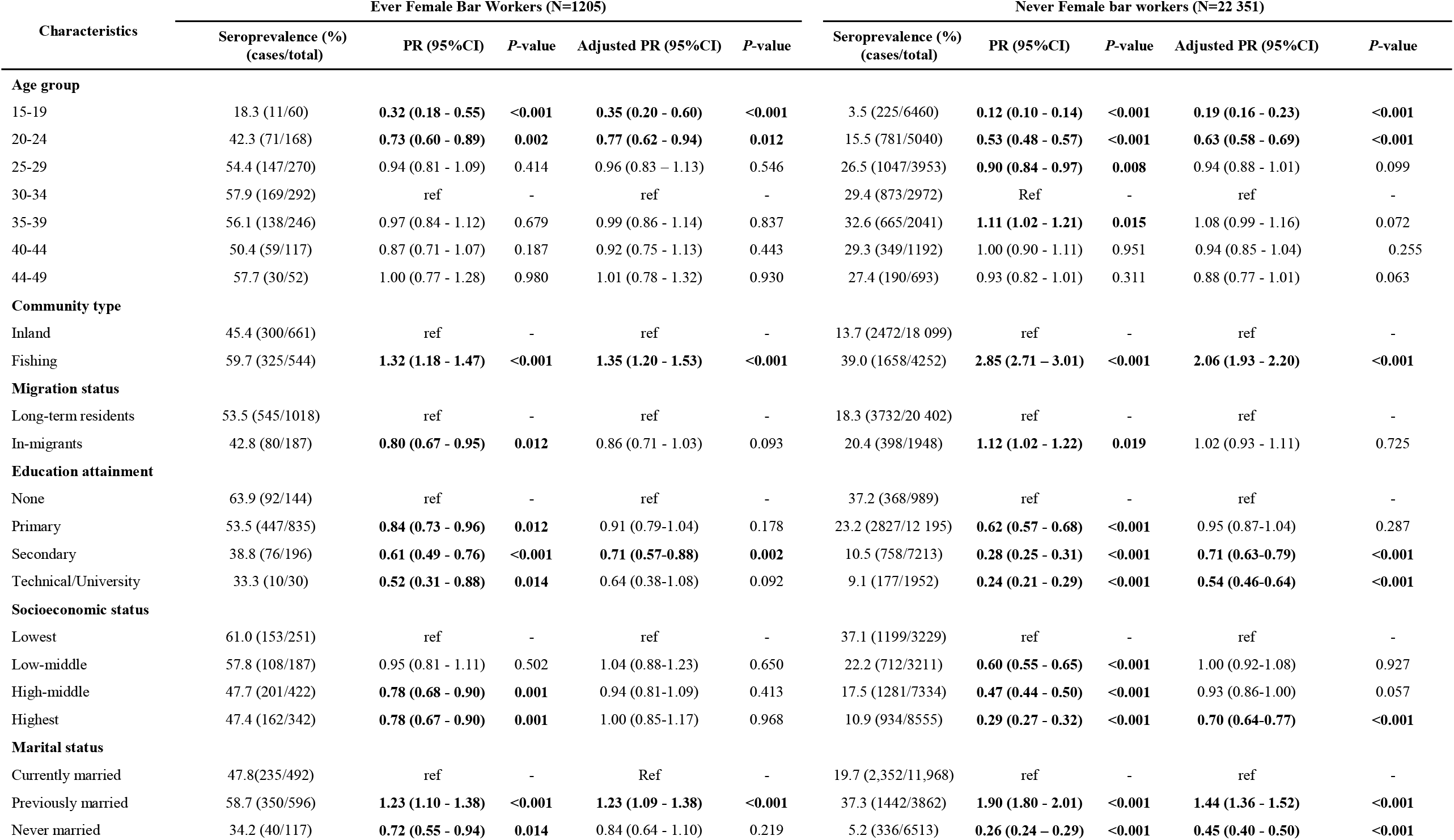
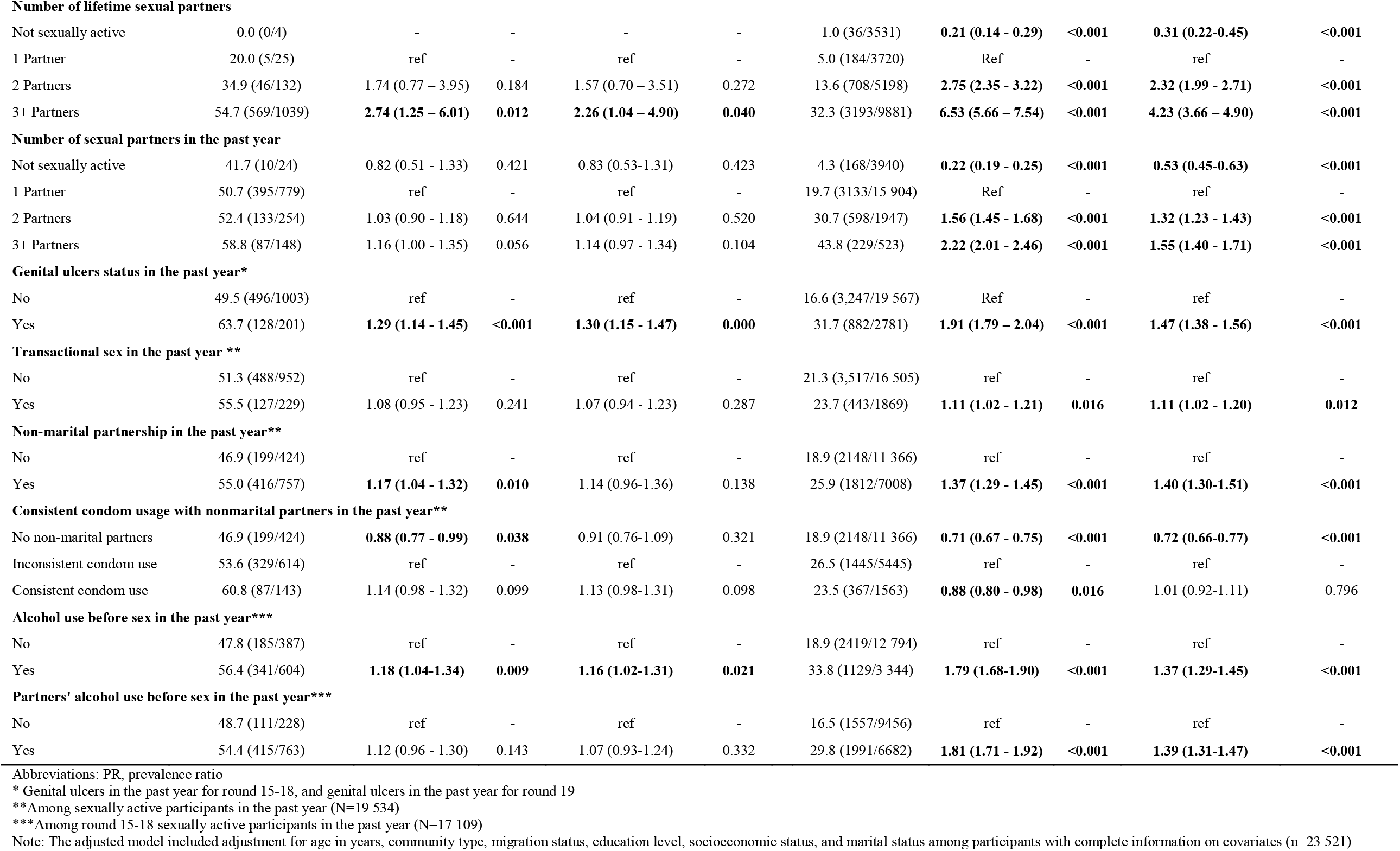
Prevalence ratios of HIV seroprevalence at baseline by self-reported history of bar work as a primary or secondary occupation among female participants in the Rakai Community Cohort Study, Uganda, 2011-2020 (N=23 556)

| Characteristics | Ever Female Bar Workers (N=1205) |  |  |  |  | Never Female bar workers (N=22 351) |  |  |  |  |
| --- | --- | --- | --- | --- | --- | --- | --- | --- | --- | --- |
|  | Seroprevalence (%)<br>(cases/total) | PR (95%CI) | P-value | Adjusted PR (95%CI) | P-value | Seroprevalence (%)<br>(cases/total) | PR (95%CI) | P-value | Adjusted PR (95%CI) | P-value |
| **Age group** |  |  |  |  |  |  |  |  |  |  |
| 15-19 | 18.3 (11/60) | **0.32 (0.18 - 0.55)** | **<0.001** | **0.35 (0.20 - 0.60)** | **<0.001** | 3.5 (225/6460) | **0.12 (0.10 - 0.14)** | **<0.001** | **0.19 (0.16 - 0.23)** | **<0.001** |
| 20-24 | 42.3 (71/168) | **0.73 (0.60 - 0.89)** | **0.002** | **0.77 (0.62 - 0.94)** | **0.012** | 15.5 (781/5040) | **0.53 (0.48 - 0.57)** | **<0.001** | **0.63 (0.58 - 0.69)** | **<0.001** |
| 25-29 | 54.4 (147/270) | 0.94 (0.81 - 1.09) | 0.414 | 0.96 (0.83 - 1.13) | 0.546 | 26.5 (1047/3953) | **0.90 (0.84 - 0.97)** | **0.008** | 0.94 (0.88 - 1.01) | 0.099 |
| 30-34 | 57.9 (169/292) | ref | - | ref | - | 29.4 (873/2972) | Ref | - | ref | - |
| 35-39 | 56.1 (138/246) | 0.97 (0.84 - 1.12) | 0.679 | 0.99 (0.86 - 1.14) | 0.837 | 32.6 (665/2041) | **1.11 (1.02 - 1.21)** | **0.015** | 1.08 (0.99 - 1.16) | 0.072 |
| 40-44 | 50.4 (59/117) | 0.87 (0.71 - 1.07) | 0.187 | 0.92 (0.75 - 1.13) | 0.443 | 29.3 (349/1192) | 1.00 (0.90 - 1.11) | 0.951 | 0.94 (0.85 - 1.04) | 0.255 |
| 44-49 | 57.7 (30/52) | 1.00 (0.77 - 1.28) | 0.980 | 1.01 (0.78 - 1.32) | 0.930 | 27.4 (190/693) | 0.93 (0.82 - 1.01) | 0.311 | 0.88 (0.77 - 1.01) | 0.063 |
| **Community type** |  |  |  |  |  |  |  |  |  |  |
| Inland | 45.4 (300/661) | ref | - | ref | - | 13.7 (2472/18 099) | ref | - | ref | - |
| Fishing | 59.7 (325/544) | **1.32 (1.18 - 1.47)** | **<0.001** | **1.35 (1.20 - 1.53)** | **<0.001** | 39.0 (1658/4252) | **2.85 (2.71 - 3.01)** | **<0.001** | **2.06 (1.93 - 2.20)** | **<0.001** |
| **Migration status** |  |  |  |  |  |  |  |  |  |  |
| Long-term residents | 53.5 (545/1018) | ref | - | ref | - | 18.3 (3732/20 402) | ref | - | ref | - |
| In-migrants | 42.8 (80/187) | **0.80 (0.67 - 0.95)** | **0.012** | 0.86 (0.71 - 1.03) | 0.093 | 20.4 (398/1948) | **1.12 (1.02 - 1.22)** | **0.019** | 1.02 (0.93 - 1.11) | 0.725 |
| **Education attainment** |  |  |  |  |  |  |  |  |  |  |
| None | 63.9 (92/144) | ref | - | ref | - | 37.2 (368/989) | ref | - | ref | - |
| Primary | 53.5 (447/835) | **0.84 (0.73 - 0.96)** | **0.012** | 0.91 (0.79-1.04) | 0.178 | 23.2 (2827/12 195) | **0.62 (0.57 - 0.68)** | **<0.001** | 0.95 (0.87-1.04) | 0.287 |
| Secondary | 38.8 (76/196) | **0.61 (0.49 - 0.76)** | **<0.001** | **0.71 (0.57-0.88)** | **0.002** | 10.5 (758/7213) | **0.28 (0.25 - 0.31)** | **<0.001** | **0.71 (0.63-0.79)** | **<0.001** |
| Technical/University | 33.3 (10/30) | **0.52 (0.31 - 0.88)** | **0.014** | 0.64 (0.38-1.08) | 0.092 | 9.1 (177/1952) | **0.24 (0.21 - 0.29)** | **<0.001** | **0.54 (0.46-0.64)** | **<0.001** |
| **Socioeconomic status** |  |  |  |  |  |  |  |  |  |  |
| Lowest | 61.0 (153/251) | ref | - | ref | - | 37.1 (1199/3229) | ref | - | ref | - |
| Low-middle | 57.8 (108/187) | 0.95 (0.81 - 1.11) | 0.502 | 1.04 (0.88-1.23) | 0.650 | 22.2 (712/3211) | **0.60 (0.55 - 0.65)** | **<0.001** | 1.00 (0.92-1.08) | 0.927 |
| High-middle | 47.7 (201/422) | **0.78 (0.68 - 0.90)** | **0.001** | 0.94 (0.81-1.09) | 0.413 | 17.5 (1281/7334) | **0.47 (0.44 - 0.50)** | **<0.001** | 0.93 (0.86-1.00) | 0.057 |
| Highest | 47.4 (162/342) | **0.78 (0.67 - 0.90)** | **0.001** | 1.00 (0.85-1.17) | 0.968 | 10.9 (934/8555) | **0.29 (0.27 - 0.32)** | **<0.001** | **0.70 (0.64-0.77)** | **<0.001** |
| **Marital status** |  |  |  |  |  |  |  |  |  |  |
| Currently married | 47.8(235/492) | ref | - | Ref | - | 19.7 (2,352/11,968) | ref | - | ref | - |
| Previously married | 58.7 (350/596) | **1.23 (1.10 - 1.38)** | **<0.001** | **1.23 (1.09 - 1.38)** | **<0.001** | 37.3 (1442/3862) | **1.90 (1.80 - 2.01)** | **<0.001** | **1.44 (1.36 - 1.52)** | **<0.001** |
| Never married | 34.2 (40/117) | **0.72 (0.55 - 0.94)** | **0.014** | 0.84 (0.64 - 1.10) | 0.219 | 5.2 (336/6513) | **0.26 (0.24 - 0.29)** | **<0.001** | **0.45 (0.40 - 0.50)** | **<0.001** |
| Number of lifetime sexual partners |  |  |  |  |  |  |  |  |  |  |
| Not sexually active | 0.0 (0/4) | - | - | - | - | 1.0 (36/3531) | **0.21 (0.14 - 0.29)** | **<0.001** | **0.31 (0.22-0.45)** | **<0.001** |
| 1 Partner | 20.0 (5/25) | ref | - | ref | - | 5.0 (184/3720) | Ref | - | ref | - |
| 2 Partners | 34.9 (46/132) | 1.74 (0.77 – 3.95) | 0.184 | 1.57 (0.70 – 3.51) | 0.272 | 13.6 (708/5198) | **2.75 (2.35 - 3.22)** | **<0.001** | **2.32 (1.99 - 2.71)** | **<0.001** |
| 3+ Partners | 54.7 (569/1039) | **2.74 (1.25 – 6.01)** | **0.012** | **2.26 (1.04 – 4.90)** | **0.040** | 32.3 (3193/9881) | **6.53 (5.66 – 7.54)** | **<0.001** | **4.23 (3.66 – 4.90)** | **<0.001** |
| Number of sexual partners in the past year |  |  |  |  |  |  |  |  |  |  |
| Not sexually active | 41.7 (10/24) | 0.82 (0.51 - 1.33) | 0.421 | 0.83 (0.53-1.31) | 0.423 | 4.3 (168/3940) | **0.22 (0.19 - 0.25)** | **<0.001** | **0.53 (0.45-0.63)** | **<0.001** |
| 1 Partner | 50.7 (395/779) | ref | - | ref | - | 19.7 (3133/15 904) | Ref | - | ref | - |
| 2 Partners | 52.4 (133/254) | 1.03 (0.90 - 1.18) | 0.644 | 1.04 (0.91 - 1.19) | 0.520 | 30.7 (598/1947) | **1.56 (1.45 - 1.68)** | **<0.001** | **1.32 (1.23 - 1.43)** | **<0.001** |
| 3+ Partners | 58.8 (87/148) | 1.16 (1.00 - 1.35) | 0.056 | 1.14 (0.97 - 1.34) | 0.104 | 43.8 (229/523) | **2.22 (2.01 - 2.46)** | **<0.001** | **1.55 (1.40 - 1.71)** | **<0.001** |
| Genital ulcers status in the past year\* |  |  |  |  |  |  |  |  |  |  |
| No | 49.5 (496/1003) | ref | - | ref | - | 16.6 (3,247/19 567) | Ref | - | ref | - |
| Yes | 63.7 (128/201) | **1.29 (1.14 - 1.45)** | **<0.001** | **1.30 (1.15 - 1.47)** | **0.000** | 31.7 (882/2781) | **1.91 (1.79 – 2.04)** | **<0.001** | **1.47 (1.38 - 1.56)** | **<0.001** |
| Transactional sex in the past year \*\* |  |  |  |  |  |  |  |  |  |  |
| No | 51.3 (488/952) | ref | - | ref | - | 21.3 (3,517/16 505) | ref | - | ref | - |
| Yes | 55.5 (127/229) | 1.08 (0.95 - 1.23) | 0.241 | 1.07 (0.94 - 1.23) | 0.287 | 23.7 (443/1869) | **1.11 (1.02 - 1.21)** | **0.016** | **1.11 (1.02 - 1.20)** | **0.012** |
| Non-marital partnership in the past year\*\* |  |  |  |  |  |  |  |  |  |  |
| No | 46.9 (199/424) | ref | - | ref | - | 18.9 (2148/11 366) | ref | - | ref | - |
| Yes | 55.0 (416/757) | **1.17 (1.04 - 1.32)** | **0.010** | 1.14 (0.96-1.36) | 0.138 | 25.9 (1812/7008) | **1.37 (1.29 - 1.45)** | **<0.001** | **1.40 (1.30-1.51)** | **<0.001** |
| Consistent condom usage with nonmarital partners in the past year\*\* |  |  |  |  |  |  |  |  |  |  |
| No non-marital partners | 46.9 (199/424) | **0.88 (0.77 - 0.99)** | **0.038** | 0.91 (0.76-1.09) | 0.321 | 18.9 (2148/11 366) | **0.71 (0.67 - 0.75)** | **<0.001** | **0.72 (0.66-0.77)** | **<0.001** |
| Inconsistent condom use | 53.6 (329/614) | ref | - | ref | - | 26.5 (1445/5445) | ref | - | ref | - |
| Consistent condom use | 60.8 (87/143) | 1.14 (0.98 - 1.32) | 0.099 | 1.13 (0.98-1.31) | 0.098 | 23.5 (367/1563) | **0.88 (0.80 - 0.98)** | **0.016** | 1.01 (0.92-1.11) | 0.796 |
| Alcohol use before sex in the past year\*\*\* |  |  |  |  |  |  |  |  |  |  |
| No | 47.8 (185/387) | ref | - | ref | - | 18.9 (2419/12 794) | ref | - | ref | - |
| Yes | 56.4 (341/604) | **1.18 (1.04-1.34)** | **0.009** | **1.16 (1.02-1.31)** | **0.021** | 33.8 (1129/3 344) | **1.79 (1.68-1.90)** | **<0.001** | **1.37 (1.29-1.45)** | **<0.001** |
| Partners' alcohol use before sex in the past year\*\*\* |  |  |  |  |  |  |  |  |  |  |
| No | 48.7 (111/228) | ref | - | ref | - | 16.5 (1557/9456) | ref | - | ref | - |
| Yes | 54.4 (415/763) | 1.12 (0.96 - 1.30) | 0.143 | 1.07 (0.93-1.24) | 0.332 | 29.8 (1991/6682) | **1.81 (1.71 - 1.92)** | **<0.001** | **1.39 (1.31-1.47)** | **<0.001** |

Abbreviations: PR, prevalence ratio
\* Genital ulcers in the past year for round 15-18, and genital ulcers in the past year for round 19
\*\*Among sexually active participants in the past year (N=19 534)
\*\*\*Among round 15-18 sexually active participants in the past year (N=17 109)
Note: The adjusted model included adjustment for age in years, community type, migration status, education level, socioeconomic status, and marital status among participants with complete information on covariates (n=23 521)

### Fluctuation in female bar worker status

Women fluctuated in and out of bar work across time. Of the 172 FBWs who completed all five surveys, only 26 (15.1%) consistently reported being a bar worker at every visit, while 59 (34.3%) indicated bar work at just a single survey (**Figure_2**). However, HIV seroprevalence appeared unrelated to the number of visits they reported bar work, or their past or future engagement in bar work (**Supplement_Figure_4**). Additionally, HIV seroprevalence was similar among FBWs regardless of whether they reported bar work as their primary occupation, secondary occupation, or both (**Supplementary_Figure_5**).

**Figure 2.**
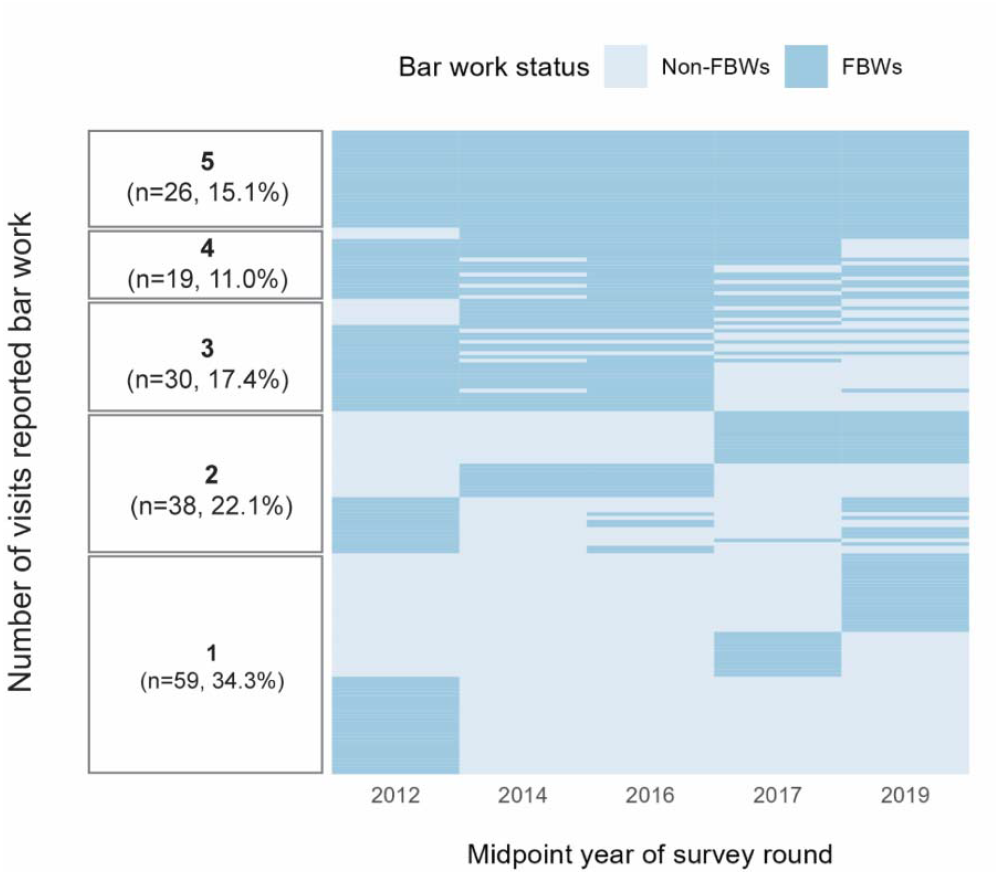
Fluctuation of bar work status by survey round among female participants with five survey visits in the Rakai Community Cohort Study, Uganda, 2011-2020 (N=172) Abbreviations: FBW, female bar worker

### HIV incidence

There were 16,142 female participants who were HIV-seronegative at their first visit; of these, 9,726 (60.3%) had at least one consecutive follow-up visit. **Supplementary_Table_4** shows the count and percentages of person visits that were either included or excluded from the incidence cohort based on participant characteristics. The proportion of excluded visits was similar among FBWs (32.7%) and non-FBWs (32.3%). There was a higher proportion of visits lost to follow-up among those aged 15-19 years, recent migrants to the study communities, those never married, and those who were not sexually active either in their lifetime or in the past year.

Overall, 356 HIV seroconversion events occurred over 39,228 person-years of follow-up. HIV seroconversion was 0.91/100 person-years, with 0.87/100 person-years among non-recent FBWs and 2.49/100 person-years among recent FBWs (IRR=3.02,95%CI=1.94-4.46; age-adjusted IRR=3.64,95%CI=2.33-5.42) (**Table_3**). Age-adjusted IRR remained similar after incorporating censoring weights (IRR=3.77,95%CI=2.41-5.61). When analyses were stratified by community type, associations between FBW status and HIV incident infection were greater in inland compared to fishing communities (3.30 vs. 1.50, *p*-interaction=0.064).

**Table 3.** Incidence rates and ratios of HIV seroconversion by the self-reported history of bar work <u>at either the start or end of a visit interval</u> among female participants in the Rakai Community Cohort Study, Uganda, 2011-2020 (N=9726)

| Population | HIV seroconversion | Person-years | IR per 100 PY (95%CI) | IRR (95%CI) | P-value | Age-adjusted IRR (95%CI) | P-value | Weighted age-adjusted IRR (95%CI) | P-value |
| --- | --- | --- | --- | --- | --- | --- | --- | --- | --- |
| **Overall** | 356 | 39 228 | 0.91 (0.82-1.01) |  |  |  |  |  |  |
| Non-FBW | 332 | 38 263 | 0.87 (0.78-0.97) | ref | - | ref | - | ref | - |
| FBW | 24 | 965 | 2.49 (1.59-3.70) | 3.02 (1.94-4.46) | <0.001 | 3.64 (2.33-5.42) | <0.001 | 3.77 (2.41-5.61) | <0.001 |
| **Inland community** | 228 | 33 777 | 0.68 (0.59-0.77) |  |  |  |  |  |  |
| Non-FBW | 215 | 33 134 | 0.65 (0.57-0.74) | ref | - | ref | - | ref | - |
| FBW | 13 | 643 | 2.02 (1.08-3.46) | 3.30 (1.79-5.54) | <0.001 | 3.97 (2.14-6.74) | <0.001 | 4.14 (2.23-7.01) | <0.001 |
| **Fishing community** | 128 | 5451 | 2.35 (1.96-2.79) |  |  |  |  |  |  |
| Non-FBW | 117 | 5129 | 2.28 (1.89-2.73) | ref | - | ref | - | ref | - |
| FBW | 11 | 322 | 3.42 (1.71-6.12) | 1.50 (0.76-2.65) | 0.199 | 1.75 (0.88-3.11) | 0.079 | 1.71 (0.86-3.06) | 0.091 |

Abbreviations: FBW, female bar worker; HIV, human immunodeficiency virus; IR, incidence rate; CI, confidence interval; IRR, incidence rate ratio; PY, person-years

**Supplementary_Table_5** shows the HIV incidence by survey period. For non-FBWs, HIV incidence declined from 1.06/100 person-years (2011-2016) to 0.41/100 person-years (2016-2020). Among FBWs, the incidence declined from 3.12/100 person-years to 1.03/100 person-years. Despite this decrease, disparities in the risk of HIV acquisition between FBWs and non-FBWs persisted with an IRR of 2.48(95%CI=0.60-6.75) during the 2016-2020 period.

In sensitivity analyses using participants’ ever or never bar work status during the five surveys as the exposure, HIV seroconversion remained higher among ever-FBWs than never-FBWs (age-adjusted IRR=3.82,95%CI=2.69-5.29) (**Supplementary_Table_6**).

### HIV viral suppression

Between 2013 and 2020 (survey rounds 16-19), 4,272 women tested HIV-seropositive and had valid viral load data available at 8,763/8,791 (99.7%) visits. The overall viral suppression (regardless of ART use) at these visits was 82.2% (6,610/8,019) among FBWs and 81.9% (591/744) among non-FBWs (age-adjusted PR=0.99, 95%CI=0.95-1.03, **Supplementary_Table_7**). HIV viral suppression significantly increased over time among both FBWs and non-FBWs (**Supplementary_Figure_6**), reaching 88.1% among FBWs and 91.0% among non-FBWs by the final survey (2018-2020).

### Population prevalence of HIV viremia

**Figure_3** shows the population prevalence and prevalence ratios of HIV viremia by FBW status. As HIV viral suppression among WLWH increased, the PPV decreased from 15.3% to 5.8% for FBWs and from 6.2% to 1.8% for non-FBWs between surveys 16 and 19. Nonetheless, the overall PPV among FBWs was 1.69 times higher than among non-FBWs across our study period, accounting for repeated measures (95%CI=1.38-2.08), which rose from 1.75(95%CI=1.33-2.32) in round 16 to 2.64(95%CI=1.68-4.17) in round 19, adjusting for age and community type. Fishing communities showed a consistently higher PPV than inland communities. However, the difference in PPV between FBWs and non-FBWs was more pronounced within inland communities (**Supplementary_Figure_7**).

**Figure 3.**
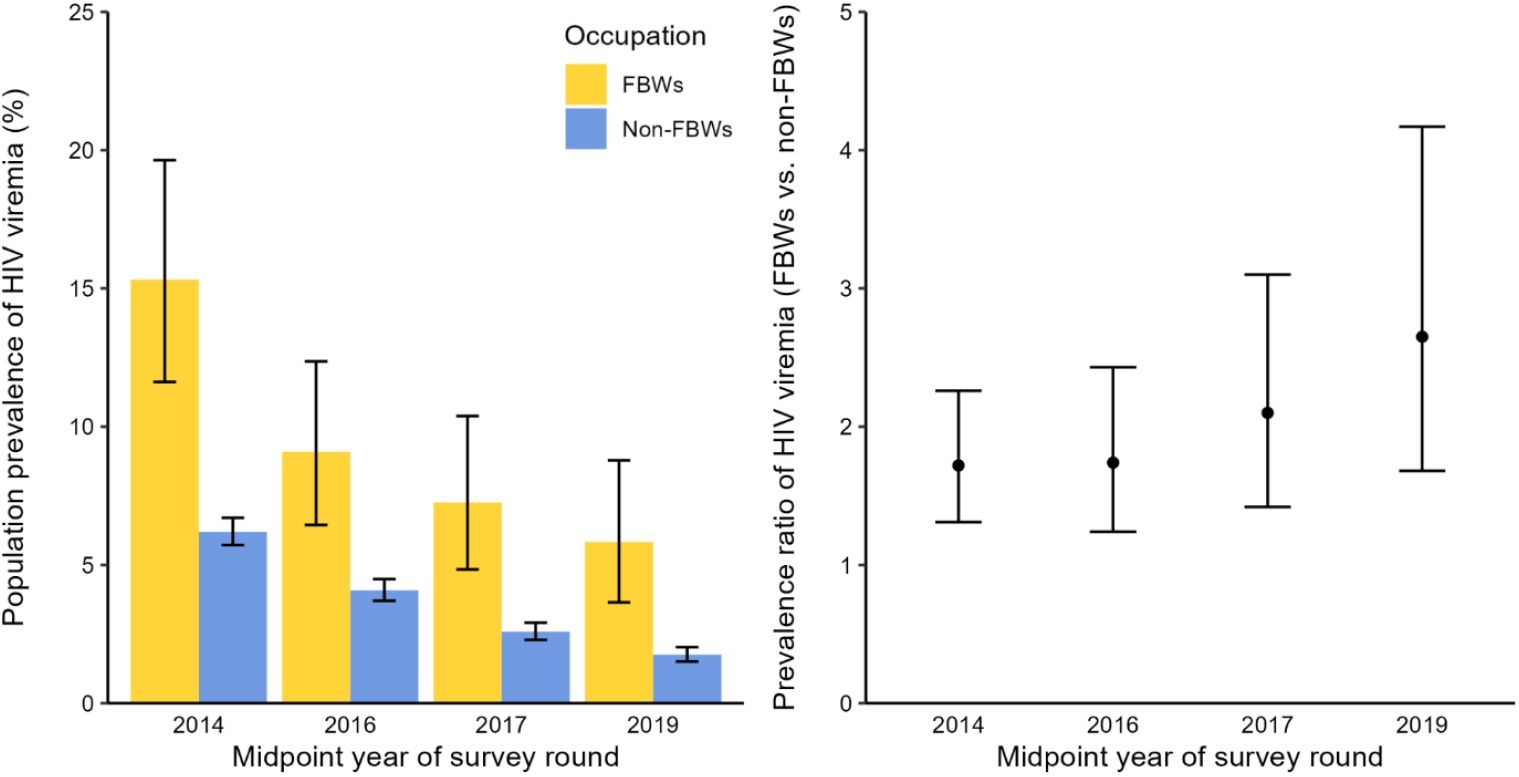
Population prevalence and prevalence ratios of HIV viremia by self-reported history of bar work as a primary or secondary occupation and by survey round among female participants of Rakai Community Cohort Study, Uganda, 2013-2020 (N=20 250) Abbreviations: FBW, female bar worker; HIV, human immunodeficiency virus Note: Prevalence ratios were comparing FBWs to non-FBWs adjusting for age and community type

### HIV awareness and PrEP use

Among the 2,135 WLWH in survey round 19 (years 2018-2020), awareness of HIV serostatus was similar between FBWs (96.6%) and non-FBWs (95.1%)(*P*=0.380)**(Supplementary_Table_8)**. Among the 7,827 HIV-seronegative women who answered questions about self-reported PrEP use in survey round 19, 79.9% (143/179) of FBWs had heard of PrEP, compared to 72.7%(5,559/7,648) of non-FBWs (p=0.032). Among FBWs, 10.6% had ever used PrEP, and 2.8% were currently using it, while among non-FBWs, 1.3% had ever and 1.4% were currently using PrEP (*P*<0.001) **(Supplementary_Table_9)**.

## Discussion

In this population-based longitudinal study of over 23,000 women, we found that FBWs had significantly higher HIV seroprevalence and incidence compared to the general female population not engaged in bar work. Despite similar sociodemographic and behavioral factors associated with prevalent HIV infections between FBWs and non-FBWs, HIV burden was consistently higher among FBWs, irrespective of self-reported behaviors or sociodemographics. This suggests that bar work, reflecting the socioeconomic conditions and/or affecting sexual behaviors tied to it, may play significant roles in the elevated HIV risk experienced by FBWs. Moreover, longitudinal data revealed that bar work was typically transient, with women already having high HIV seroprevalence prior to their first report of bar work. Taken together with data on higher HIV incidence among FBWs, our results suggest that infection risk likely precedes and persists throughout their time in the occupation. Our findings also highlight that few HIV-seronegative FBWs reported using PrEP despite high PrEP awareness. Furthermore, although FBWs and non-FBWs living with HIV had similar levels of HIV serostatus awareness and viral suppression, the population prevalence of HIV viremia was substantially higher among FBWs due to their elevated background disease burden. In summary, our results underscore the need for targeted HIV prevention strategies for FBWs and women at risk of entry into bar work.

The substantially higher HIV seroprevalence among FBWs in this study aligns limited data from other African settings. Notably, HIV seroprevalence among FBWs in our study population (51.9%) was higher than the 31.4% seroprevalence reported among FSWs in the 2021 CRANE survey in Uganda.^22^ Beyond sex work, prior qualitative research suggests that elevated HIV burden in FBWs may be attributed to factors such as low SES, low educational attainment, and social marginalization resulting from partner separation and divorce, which was consistent with our findings.^15,23^

Although only one-fifth of FBWs reported engaging in transactional sex in our study, prior qualitative research conducted in the same communities as this study and elsewhere suggest that female bar and sex work are deeply intertwined.^5,7^ Indeed, HIV incidence rates among FBWs in this study, estimated at 2.51/100 person-years overall and 3.42/100 person-years in fishing communities, are on par with that in studies among African FSWs, including in Uganda.^24^ PrEP has already been shown to be highly effective in preventing HIV among FSWs and would likely benefit FBWs as well.^25^ However, despite high awareness of PrEP among FBWs in our study, only 13.4% had ever used it and <3% were currently using it.

Despite similarly high proportions of viral suppression (>80%) between FBWs and non-FBWs living with HIV, the population prevalence of viremia was significantly higher among FBWs. This aligns with other studies of African women, including FSWs, which show comparable viral suppression rates among those on ART.^26,27^ However, the higher burden of viremia among FBWs indicates that although treatment programs are effective in achieving suppression in those with HIV, the high background HIV acquisition risk in this population may lead to increased risk for onward HIV transmission to their partners. Overall, these data underscore the need to prioritize HIV testing and prevention interventions for FBWs.

The transient nature of bar work was another notable finding. FBWs frequently moved in and out of the occupation and were more likely to report recent migration. Prior literature indicates that both bar work and sex work are characterized by high mobility and short-term engagement.^28^ Qualitative studies suggest that this transience is partly due to a similarly transient client population at bars and temporary needs for financial support.^29,30^ Such mobility may pose significant challenges for the sustained delivery of HIV prevention and treatment services. Implementation studies are needed to assess how best to deliver effective interventions, ensuring that HIV services are accessible and adaptable for this population.

This study had several limitations. First, the reliance on self-reported occupation data may introduce bias, as bar work could be underreported due to stigma similar to that faced by sex workers. Second, we did not assess specific occupational exposures, making it difficult to determine whether the elevated HIV risk among FBWs was directly related to bar work or to other risk factors (e.g., lower SES). Finally, while our study spanned nearly a decade and included a large sample size with a high participation rate, data were collected exclusively in southern Uganda and may not be fully representative of the broader African HIV epidemic.

In conclusion, FBWs in Uganda have higher HIV seroprevalence, increased risk of HIV acquisition, and greater population prevalence of HIV viremia than non-FBWs. Given the heightened HIV risk and low uptake of preventive measures among FBWs, our findings underscore the importance of prioritizing FBWs in HIV control efforts. Tailored interventions addressing their unique challenges might be particularly beneficial if focused on women prior to or immediately after entering bar work, potentially mitigating HIV risk for both FBWs and their sexual partners.

## Data Availability

All data produced in the present study are available upon reasonable request to the Rakai Health Sciences Program.

## Contributors

XF, EUP, CEK, AART, and MKG conceived and designed the study. GN, FN, GK, RS, HN, GK, and RMG collected and managed the data. XF and SS conducted statistical methods and analyses. All authors contributed to the interpretation of the findings. XF, CEK, AART, and MKG drafted the manuscript, and all authors participated in revising it. All authors had access to all of the data in the study and read and approved the final manuscript for publication.

## Declaration of interests

We declare no competing interests.

## Data sharing

De-identified Rakai Community Cohort Study data can be provided to interested parties subject to the completion of the Rakai Health Sciences Program data request form and the signing of a Data Transfer Agreement. Inquiries should be directed to.

## Acknowledgment

We sincerely thank all RCCS participants for their time and valuable contributions to this study. We also acknowledge the dedication and hard work of the data collection, laboratory, service linkage, and data management staff at RHSP. We thank members of the Johns Hopkins Bloomberg School of Public Health and NIAID Laboratory of Immunoregulation International HIV/STD Section at the Johns Hopkins University School of Medicine for their helpful contributions to this work. This study was supported in part by the National Institutes of Health (NIH) National Institute of Allergy and Infectious Diseases (NIAID) (U01AI075115, R01AI087409, U01AI100031, R01AI110324, R01AI114438, K25AI114461, R01AI123002, R01AI128779, R01AI143333, R21AI145682, R01AI155080, ZIAAI001040), NIH National Institute of Child Health and Development (R01HD050180, R01HD070769, R01HD091003), NIH National Heart, Lung, and Blood Institute (R01HL152813), National Institute of Diabetes, Digestive, and Kidney Diseases (R01DK131926), the Fogarty International Center (D43TW009578, D43TW010557), the Johns Hopkins University Center for AIDS Research (P30AI094189). This project was supported by the National Institutes of Health and in part by the President’s Emergency Plan for AIDS Relief (PEPFAR) through the U.S. Centers for Disease Control and Prevention (U.S. CDC) cooperative agreement number NU2GGH002009 and NU2GGH000817. This project was also supported in part by the Division of Intramural Research, NIAID, NIH.

## Notes

### Competing Interest Statement

The authors have declared no competing interest.

### Author Declarations

Ethics committee/IRB of Uganda Virus Research Institute gave ethical approval for this work. Ethics committee/IRB of Johns Hopkins School of Medicine gave ethical approval for this work.

