## Supplementary Materials for "HIV seroprevalence, incidence, and viral suppression among Ugandan female bar workers: a population-based study"

### **Supplementary Statistical Methods**

All statistical analyses were conducted using the Stata 15.1 and R statistical software (version 4.2.1).

#### Imputation of missing occupation from survey round 16

Of the 23,556 participants, 66.8% reported a single occupation during the study period, and 25.0% reported two different occupations. We imputed the occupation for 2609 participants who were not surveyed about their occupation in survey round 16, using the nearest reported occupation from other rounds. Among these, the 116 participants who only participated in survey round 16 were classified as non-FBWs, and there were 19.2%, 78.8%, 1.5%, and 0.4% of the 2493 missing occupation data were imputed from survey rounds 15, 17, 18, and 19, respectively. Finally, 37 participants were imputed as FBWs, and 2572 were imputed as non-FBWs in survey round 16. The proportion of FBWs in survey round 16 was 3.2% (223/7060) before imputation and 2.7% (260/9669) afterward.

| **Number of occupations reported during our study period** | **Count** | **Percent** |
| --- | --- | --- |
| 1 | 15733 | 66.8% |
| 2 | 5882 | 25.0% |
| 3 | 1731 | 7.3% |
| 4 | 206 | 0.9% |
| 5 | 4 | 0.0% |

| **The source survey round for the imputed occupation in round 16** | **Count** | **Percent** |
| --- | --- | --- |
| 15 | 478 | 19.2% |
| 17 | 1,966 | 78.8% |
| 18 | 38 | 1.5% |
| 19 | 11 | 0.4% |

#### Calculation on inverse probability of censoring weight (IPCW)

For each participant, logistic regression was used to estimate the probability of follow-up, considering factors including age, community type, migration status, education level, SES, marital status, number of lifetime and past-year sexual partners. We assigned each individual a weight of 1/probability and stabilized these weights, ensuring a mean weight of 1 in our study population. The IPCW was then incorporated into the Poisson regressions to calculate incidence rate ratios of HIV infection between FBWs and non-FBWs, accounting for selection bias due to differences between participants observed in the incidence cohort and those lost to follow-up.

### **Supplementary Tables and Figures**

#### Supplementary Table 1. Participation rates by survey round of the Rakai Community Cohort Study, Uganda, 2011-2020 (N=23 556)

| **Survey round** | **Number of participants** | **Number of eligible women** | **Participation rate (%)** |
| --- | --- | --- | --- |
| 15 | 9868 | 14 822 | 66.6 |
| 16 | 9669 | 15 140 | 63.9 |
| 17 | 10 441 | 16 377 | 63.8 |
| 18 | 10 540 | 16 636 | 63.4 |
| 19 | 10 414 | 17 147 | 60.7 |
| Overall | 23 556 | 36 268 | 64.9 |

Note: participation rates were calculated as the proportion of participants among women eligible in the community at the time of the survey

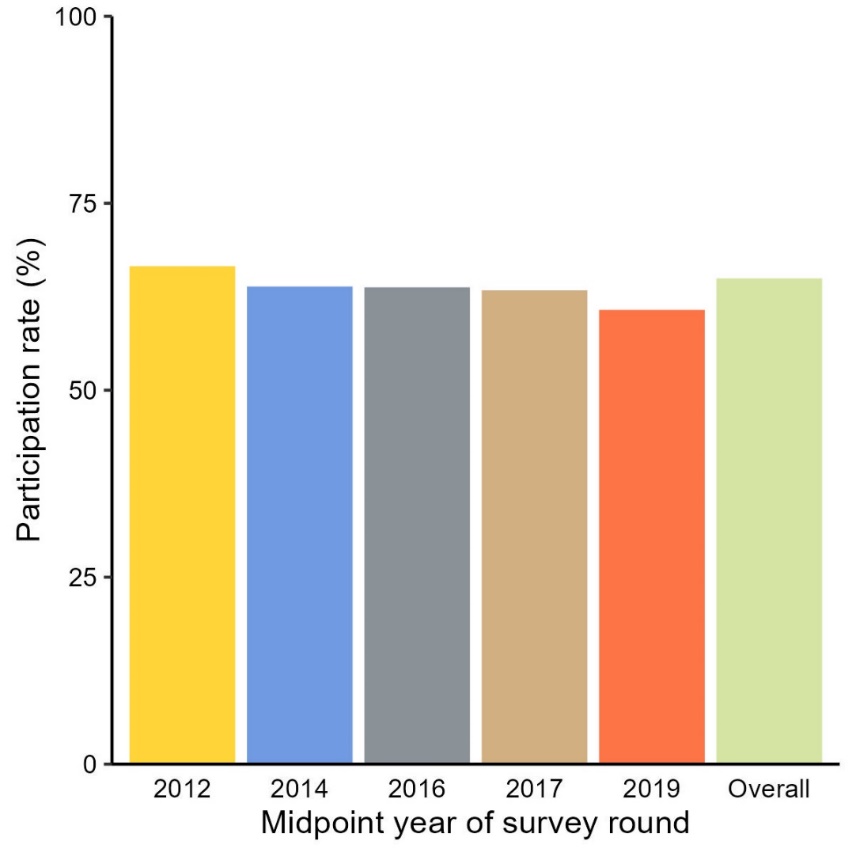

#### Supplementary Figure 1. Participation rates by survey round of the Rakai Community Cohort Study, Uganda, 2011-2020 (N=23 556)

#### Supplementary Table 2. HIV seroprevalences and prevalence ratios at baseline by self-reported history of bar work as a primary or secondary occupation among female participants in the Rakai Community Cohort Study, Uganda, 2011-2020 (N=23 556)

| **Population** | **Seroprevalence (case/total)** | **Model 1** | | **Model 2** | | **Model 3** | | **Model 4** | |
| --- | --- | --- | --- | --- | --- | --- | --- | --- | --- |
|  |  | **PR**  **(95%CI)** | ***P-*value** | **Adjusted PR (95%CI)** | ***P-*value** | **Adjusted PR (95%CI)** | ***P-*value** | **Adjusted PR (95%CI)** | ***P-*value** |
| **Overall** |  |  |  |  |  |  |  |  |  |
| Non-FBWs | 18.5% (4130/22 351) | ref |  | ref |  | ref |  | ref |  |
| FBWs | 51.9% (625/1205) | 2.81 (2.64-2.98) | <0.001 | 2.17 (2.03-2.31) | <0.001 | 1.56 (1.46-1.66) | <0.001 | 1.37 (1.28-1.46) | <0.001 |
| **Inland community** |  |  |  |  |  |  |  |  |  |
| Non-FBWs | 13.7% (2472/18 099) | ref |  | ref |  | ref |  | ref |  |
| FBWs | 45.4% (300/661) | 3.32 (3.03-3.64) | <0.001 | 2.27 (2.06-2.51) | <0.001 | 1.85 (1.68-2.04) | <0.001 | 1.55 (1.41-1.71) | <0.001 |
| **Fishing community** |  |  |  |  |  |  |  |  |  |
| Non-FBWs | 39.0% (1658/4252) | ref |  | ref |  | ref |  | ref |  |
| FBWs | 59.7% (325/544) | 1.53 (1.42-1.66) | <0.001 | 1.39 (1.28-1.50) | <0.001 | 1.40 (1.29-1.52) | <0.001 | 1.25 (1.16-1.36) | <0.001 |

Abbreviation: FBW, female bar worker; PR, prevalence ratio

Note: **Model 1** provided unadjusted results; **Model 2** adjusted for age in years; **Model 3** additionally adjusted for (community type), migration status, education attainment, socioeconomic status, and marital status among participants with complete data on covariates (n=23 521); **Model 4** additionally adjusted for the number of lifetime sexual partners at the time of visit, and the past-year self-reported number of sexual partners, genital ulcer (GUD) status, transactional sex, non-marital partnerships, and consistent condom usage with non-marital partners among participants from survey rounds 15-19 who were sexually active in the past year and had complete data on covariates (n=19 509).

#### Supplementary Table 3. HIV seroprevalences and prevalence ratios at baseline by self-reported history of bar work as a primary or secondary occupation among female participants in the Rakai Community Cohort Study, Uganda, 2011-2020 (N=23 556)

| **Characteristics** | **HIV Seroprevalence (%) (cases/total)** | | **PR (95%CI)** | ***P*-value** | **Adjusted PR (95%CI)** | ***P*-value** |
| --- | --- | --- | --- | --- | --- | --- |
|  | **Female Bar Worker** | **Non-Female bar worker** |  |  |  |  |
| **Age group** |  |  |  |  |  |  |
| 15-19 | 18.3 (11/60) | 3.5 (225/6460) | 5.26 (3.04 - 9.12) | <0.001 | 2.32 (1.28 - 4.21) | 0.006 |
| 20-24 | 42.3 (71/168) | 15.5 (781/5040) | 2.73 (2.26 - 3.29) | <0.001 | 1.67 (1.35 - 2.07) | <0.001 |
| 25-29 | 54.4 (147/270) | 26.5 (1,047/3953) | 2.06 (1.82 - 2.32) | <0.001 | 1.43 (1.25 - 1.63) | <0.001 |
| 30-34 | 58.1 (169/291) | 29.4 (873/2972) | 1.97 (1.76 - 2.21) | <0.001 | 1.42 (1.26 - 1.60) | <0.001 |
| 35-39 | 56.1 (138/246) | 32.6 (665/2041) | 1.72 (1.52 - 1.95) | <0.001 | 1.35 (1.19 - 1.53) | <0.001 |
| 40-44 | 49.6 (58/117) | 29.3 (349/1192) | 1.72 (1.41 - 2.10) | <0.001 | 1.62 (1.31 - 1.98) | <0.001 |
| 44-49 | 58.5 (31/53) | 27.4 (190/693) | 2.10 (1.62 - 2.74) | <0.001 | 1.95 (1.46 - 2.62) | <0.001 |
| **Community type** |  |  |  |  |  |  |
| Inland | 45.4 (300/661) | 13.7 (2472/18 099) | 3.32 (3.03 - 3.64) | <0.001 | 1.56 (1.42 - 1.72) | <0.001 |
| Fishing | 59.7 (325/544) | 39.0 (1658/4252) | 1.53 (1.42 - 1.66) | <0.001 | 1.26 (1.16 - 1.36) | <0.001 |
| **Migration status** |  |  |  |  |  |  |
| Long-term residents | 53.5 (545/1018) | 18.3 (3732/20 402) | 2.93 (2.74 - 3.12) | <0.001 | 1.56 (1.46 - 1.67) | <0.001 |
| In-migrants | 42.8 (80/187) | 20.4 (398/1948) | 2.09 (1.74 - 2.53) | <0.001 | 1.52 (1.24 - 1.85) | <0.001 |
| **Education attainment** |  |  |  |  |  |  |
| None | 63.9 (92/144) | 37.2 (368/989) | 1.72 (1.48 - 1.99) | <0.001 | 1.48 (1.26 - 1.73) | <0.001 |
| Primary | 53.5 (447/835) | 23.2 (2827/12 195) | 2.31 (2.15 - 2.48) | <0.001 | 1.56 (1.44 - 1.68) | <0.001 |
| Secondary | 38.8 (76/196) | 10.5 (758/7213) | 3.69 (3.06 - 4.45) | <0.001 | 1.62 (1.34 - 1.97) | <0.001 |
| Technical/University | 33.3 (10/30) | 9.1 (177/1952) | 3.68 (2.17 - 6.22) | <0.001 | 2.49 (1.44 - 4.32) | 0.001 |
| **Socioeconomic status** |  |  |  |  |  |  |
| Lowest | 61.0 (153/251) | 37.1 (1199/3229) | 1.64 (1.47 - 1.83) | <0.001 | 1.31 (1.17 - 1.46) | <0.001 |
| Low-middle | 57.8 (108/187) | 22.2 (712/3211) | 2.60 (2.27- 2.99) | <0.001 | 1.54 (1.32 - 1.79) | <0.001 |
| High-middle | 47.7 (201/422) | 17.5 (1281/7334) | 2.73 (2.44 - 3.05) | <0.001 | 1.51 (1.34 - 1.71) | <0.001 |
| Highest | 47.4 (162/342) | 10.9 (934/8555) | 4.34 (3.82 - 4.93) | <0.001 | 1.97 (1.72 - 2.27) | <0.001 |
| **Marital status** |  |  |  |  |  |  |
| Currently married | 47.8(235/492) | 19.7 (2,352/11,968) | 2.43 (2.20 - 2.68) | <0.001 | 1.63 (1.47 - 1.80) | <0.001 |
| Previously married | 58.7 (350/596) | 37.3 (1442/3862) | 1.57 (1.45 - 1.70) | <0.001 | 1.46 (1.34 - 1.58) | <0.001 |
| Never married | 34.2 (40/117) | 5.2 (336/6513) | 6.63 (5.05 - 8.70) | <0.001 | 1.96 (1.37 - 2.80) | <0.001 |
| **Number of lifetime sexual partners** | | | | | | |
| Not sexually active | 0.0 (0/4) | 1.0 (36/3531) | - | - | - | - |
| 1 Partner | 20.0 (5/25) | 5.0 (184/3720) | 4.04 (1.82 - 8.97) | 0.001 | 2.85 (1.24 - 6.58) | 0.014 |
| 2 Partners | 34.9 (46/132) | 13.6 (708/5198) | 2.56 (2.01 - 3.26) | <0.001 | 1.82 (1.43 - 2.31) | <0.001 |
| 3+ Partners | 54.7 (569/1039) | 32.3 (3193/9881) | 1.69 (1.59 - 1.80) | <0.001 | 1.34 (1.26 - 1.43) | <0.001 |
| **Number of sexual partners in the past year** | | | | | | |
| Not sexually active | 41.7 (10/24) | 4.3 (168/3940) | 9.77 (5.95 - 16.05) | <0.001 | 1.73 (1.04 – 2.86) | 0.022 |
| 1 Partner | 50.7 (395/779) | 19.7 (3133/15 904) | 2.57 (2.38 - 2.78) | <0.001 | 1.60 (1.47 - 1.73) | <0.001 |
| 2 Partners | 52.4 (133/254) | 30.7 (598/1947) | 1.70 (1.49 - 1.95) | <0.001 | 1.37 (1.19 - 1.57) | <0.001 |
| 3+ Partners | 58.8 (87/148) | 43.8 (229/523) | 1.34 (1.14 - 1.59) | 0.001 | 1.20 (1.00 - 1.42) | 0.044 |
| **Genital ulcers status in the past year*** | | | | | | |
| No | 49.5 (496/1003) | 16.6 (3,247/19 567) | 2.98 (2.78 – 3.20) | <0.001 | 1.58 (1.46 - 1.70) | <0.001 |
| Yes | 63.7 (128/201) | 31.7 (882/2781) | 2.01 (1.78 – 2.26) | <0.001 | 1.49 (1.33 - 1.68) | <0.001 |
| **Transactional sex in the past year **** | | | | | | |
| No | 51.3 (488/952) | 21.3 (3,517/16 505) | 2.41 (2.25 - 2.58) | <0.001 | 1.54 (1.43 - 1.66) | <0.001 |
| Yes | 55.5 (127/229) | 23.7 (443/1869) | 2.34 (2.03 - 2.70) | <0.001 | 1.50 (1.29 - 1.75) | <0.001 |
| **Non-marital partnership in the past year**** | | | | | | |
| No | 46.9 (199/424) | 18.9 (2148/11 366) | 2.48 (2.23 - 2.77) | <0.001 | 1.68 (1.49 - 1.87) | <0.001 |
| Yes | 55.0 (416/757) | 25.9 (1812/7008) | 2.13 (1.97 - 2.29) | <0.001 | 1.45 (1.34 - 1.56) | <0.001 |
| **Consistent condom usage with nonmarital partners in the past year**** | | | | | | |
| No non-marital partners | 46.9 (199/424) | 18.9 (2148/11 366) | 2.48 (2.23 - 2.77) | <0.001 | 1.68 (1.50 - 1.87) | <0.001 |
| Inconsistent condom use | 53.6 (329/614) | 26.5 (1445/5445) | 2.02 (1.85 - 2.20) | <0.001 | 1.44 (1.32 - 1.58) | <0.001 |
| Consistent condom use | 60.8 (87/143) | 23.5 (367/1563) | 2.59 (2.21 - 3.04) | <0.001 | 1.44 (1.21 - 1.70) | <0.001 |
| **Alcohol use before sex in the past year***** | | | | | | |
| No | 47.8 (185/387) | 18.9 (2419/12 794) | 2.53 (2.26 - 2.82) | <0.001 | 1.53 (1.36 - 1.72) | <0.001 |
| Yes | 56.4 (341/604) | 33.8 (1129/3344) | 1.67 (1.53 - 1.82) | <0.001 | 1.38 (1.26 - 1.50) | <0.001 |
| **Partners' alcohol use before sex in the past year***** | | | | | | |
| No | 48.7 (111/228) | 16.5 (1557/9456) | 2.96 (2.57 - 3.40) | <0.001 | 1.70 (1.46 - 1.97) | <0.001 |
| Yes | 54.4 (415/763) | 29.8 (1991/6682) | 1.83 (1.69 - 1.97) | <0.001 | 1.40 (1.30 - 1.52) | <0.001 |

Abbreviations: PR, prevalence ratio

* Genital ulcers in the past year for round 15-18, and genital ulcers in the past year for round 19
**Among sexually active participants in the past year (N=19 534)
***Among round 15-18 sexually active participants in the past year (N=17 109)

Note: The adjusted model included adjustment for age in years, community type, migration status, education level, socioeconomic status, and marital status among participants with complete information on covariates (n=23 521).

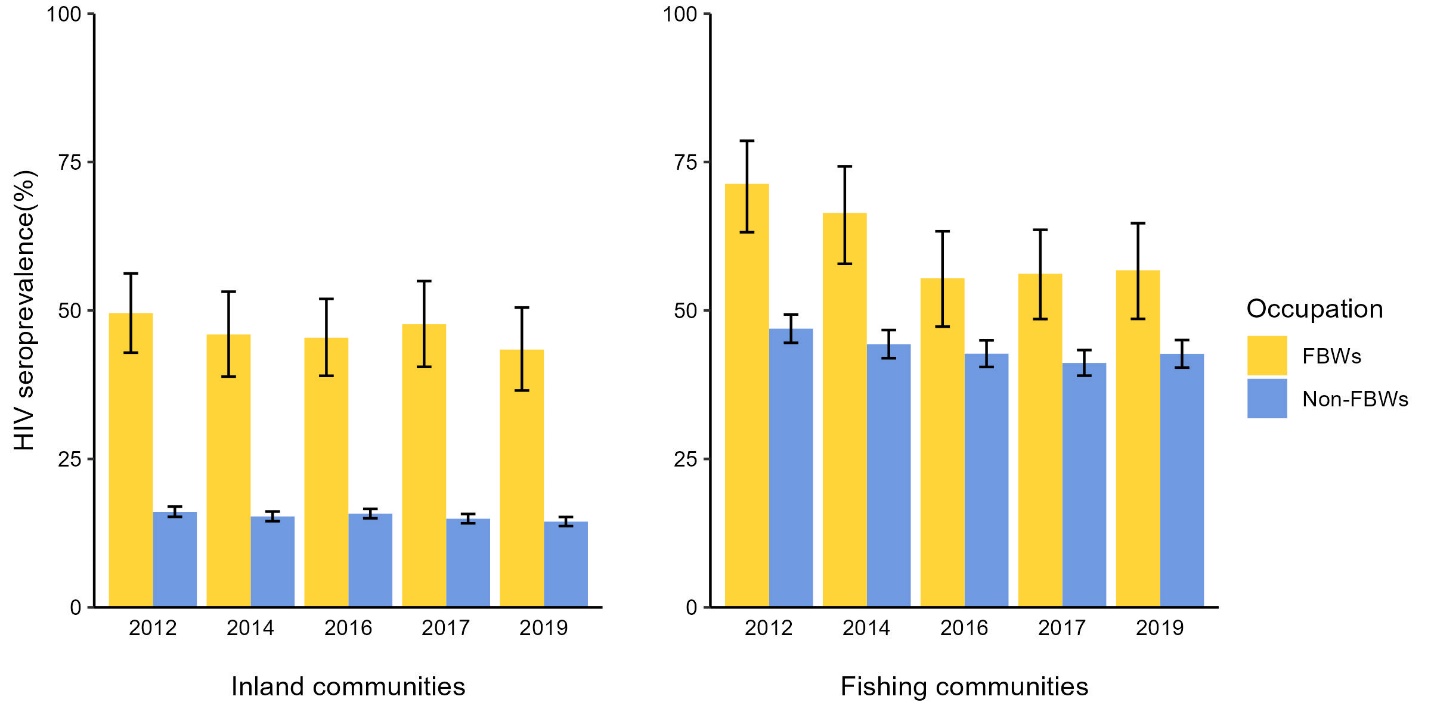

Abbreviations: FBW, female bar worker; HIV, human immunodeficiency virus

#### Supplementary Figure 2. HIV seroprevalence by self-reported history of bar work as a primary or secondary occupation and by survey round in the inland and fishing communities among female participants of Rakai Community Cohort Study, Uganda, 2011-2020 (N=23 556)

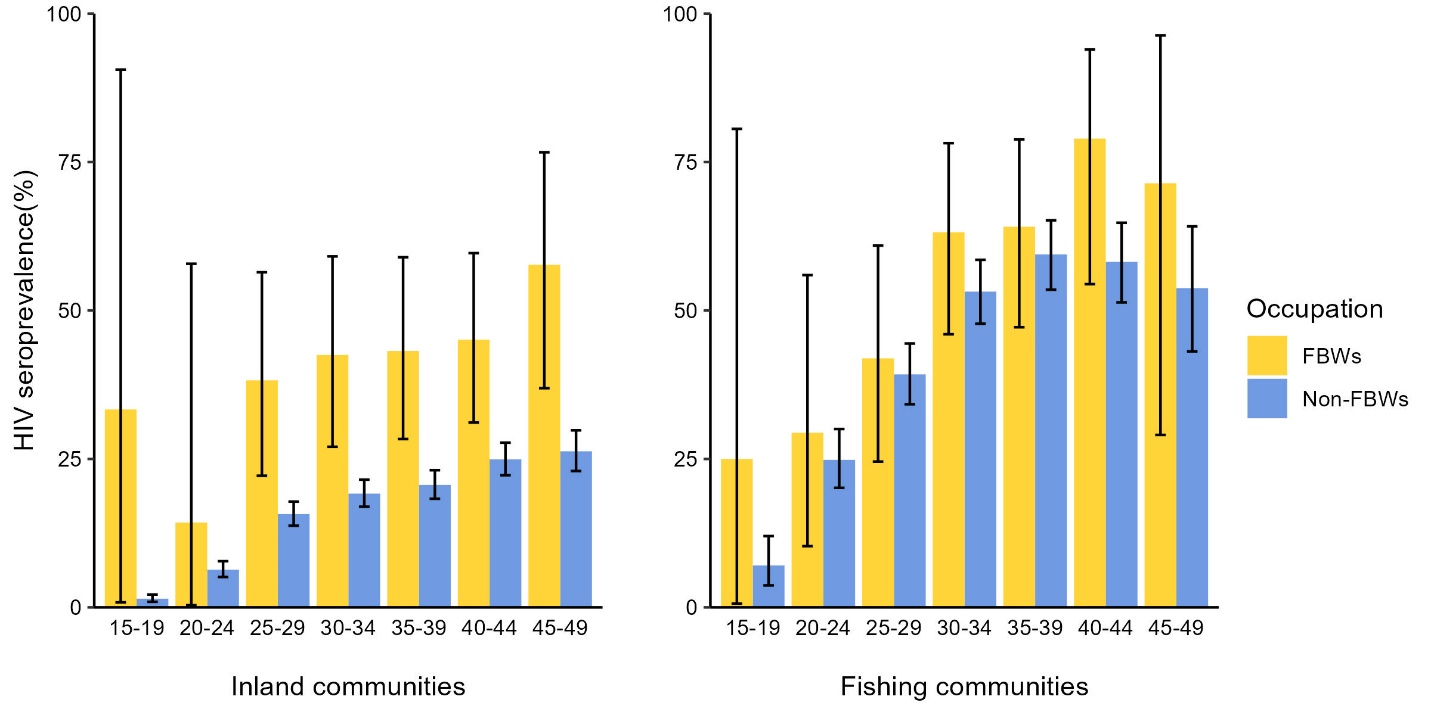

Abbreviations: FBW, female bar worker; HIV, human immunodeficiency virus

#### Supplementary Figure 3. HIV seroprevalence by self-reported history of bar work as a primary or secondary occupation in the inland and fishing communities among female participants of the survey round 19 of Rakai Community Cohort Study, Uganda, 2018-2020 (N=10 414)

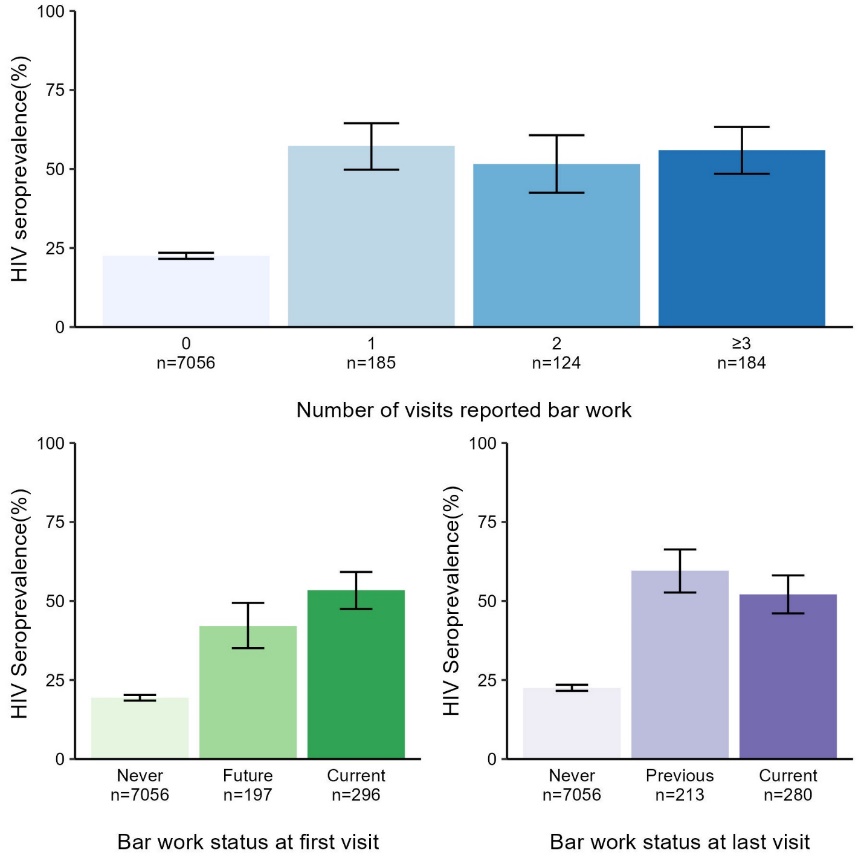

Abbreviations: HIV, human immunodeficiency virus

#### Supplementary Figure 4. HIV seroprevalence among female participants with ≥3 visits in the Rakai Community Cohort Study, Uganda, 2012-2019 (N=7549)

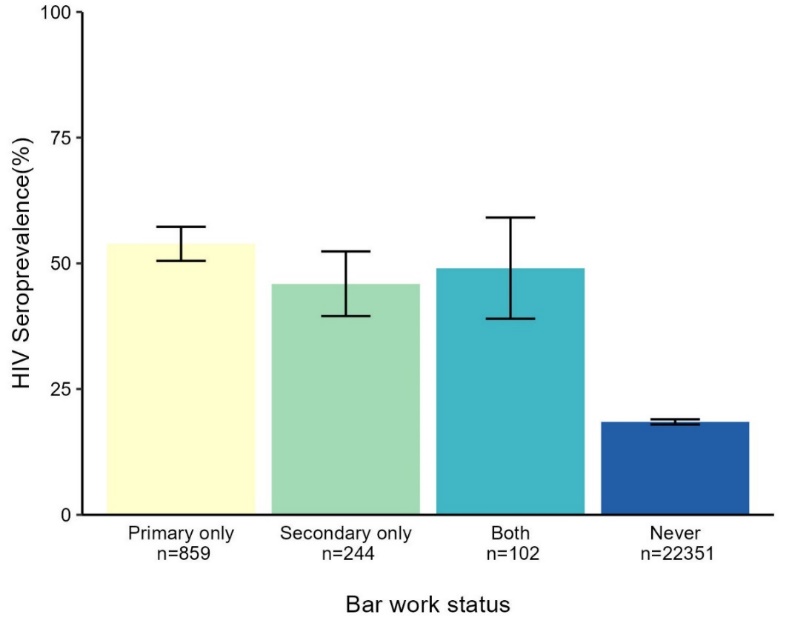

Abbreviations: HIV, human immunodeficiency virus

#### Supplementary Figure 5. HIV seroprevalence by self-reported history of bar work as a primary or secondary occupation among female participants in the Rakai Community Cohort Study, Uganda, 2011-2020 (N=23 556)

#### Supplementary Table 4. Characteristics of individual visits included in the incidence cohort and excluded due to loss to follow-up among female participants in the Rakai Community Cohort Study, Uganda, 2011-2020 (N _visit_ = 32 508)

| **Characteristics** | **Included visits** | **Excluded visits** | ***P-*value** |
| --- | --- | --- | --- |
|  | **N _visit_ = 22 009** | **N _visit_ = 10 499** |  |
| **Bar work** |  |  | 0.780 |
| No | 21 438 (67.7%) | 10 221 (32.3%) |  |
| Yes | 571 (67.3%) | 278 (32.7%) |  |
| **Age category** |  |  | <0.001 |
| 15-19 | 3484 (51.7%) | 3258 (48.3%) |  |
| 20-24 | 3838 (59.2%) | 2646 (40.8%) |  |
| 25-29 | 4048 (70.6%) | 4683 (29.4%) |  |
| 30-34 | 4103 (79.3%) | 1072 (20.7%) |  |
| 35-39 | 3327 (81.7%) | 747 (18.3%) |  |
| 40-44 | 2296 (83.9%) | 440 (16.1%) |  |
| 44-49 | 913 (58.3%) | 653 (41.7%) |  |
| **Community type** |  |  | <0.037 |
| Inland | 18 289(67.5%) | 8821 (32.5%) |  |
| Fishing | 3720 (68.9%) | 1678 (31.1%) |  |
| **Migration** |  |  | <0.001 |
| Long-term resident | 21 492 (69.2%) | 9556 (30.8%) |  |
| In-migrants | 517 (35.4%) | 943 (64.6%) |  |
| **Education attainment** | |  | <0.001 |
| None | 929 (69.9%) | 400 (30.1%) |  |
| Primary | 12 768 (69.9%) | 5511 (30.1%) |  |
| Secondary | 6728 (63.9%) | 3795 (36.1%) |  |
| Technical/University | 1576 (66.5%) | 793 (33.5%) |  |
| Missing | 8 (100.0%) | 0 (0.0%) |  |
| **Socioeconomic status** | |  | <0.001 |
| Lowest | 2735 (70.2%) | 1164 (29.9%) |  |
| Low-middle | 3140 (69.8%) | 1356 (30.2%) |  |
| High-middle | 4721 (65.8%) | 3492 (34.2%) |  |
| Highest | 9385 (67.7%) | 4473 (32.3%) |  |
| Missing | 28 (66.7%) | 14 (33.3%) |  |
| **Marital status** |  |  | <0.001 |
| Currently married | 14 658 (74.0%) | 5164 (26.1%) |  |
| Previously married | 3290 (66.7%) | 1650 (33.4%) |  |
| Never married | 4055 (52.4%) | 3681 (47.6%) |  |
| Missing | 6 (60.0%) | 4 (40.0%) |  |
| **Number of lifetime sexual partners** | |  | <0.001 |
| Not sexually active | 1938 (53.3%) | 1701 (46.7%) |  |
| 1 Partner | 4464 (67.7%) | 2135 (32.4%) |  |
| 2 Partners | 5966 (70.6%) | 2485 (29.4%) |  |
| 3+ Partners | 9630 (69.8%) | 4176 (30.3%) |  |
| Missing | 11 (84.6%) | 2 (15.4%) |  |
| **Number of sexual partners in the past year** | | | <0.001 |
| Not sexually active | 2828 (57.5%) | 2091 (42.5%) |  |
| 1 Partner | 17 444 (70.4%) | 7336 (29.6%) |  |
| 2 Partners | 1289 (60.9%) | 828 (39.1%) |  |
| 3+ Partners | 208 (51.6%) | 195 (48.4%) |  |
| Missing | 240 (83.0%) | 49 (17.0%) |  |

#### Supplementary Table 5. Incidence rates and ratios of HIV seroconversion by the self-reported history of bar work at either the start or end of a visit interval among female participants in the Rakai Community Cohort Study, Uganda, 2011-2020 (N=9726)

| **Survey round** | **HIV seroconversion** | **Person-years** | **IR per 100 PY (95%CI)** | **IRR (95% CI)** | ***P*-value** | **Age-adjusted IRR (95%CI)** | ***P*-value** |
| --- | --- | --- | --- | --- | --- | --- | --- |
| **Round 15-17** | 305 | 27 352 | 1.12 (0.99-1.25) |  |  |  |  |
| Non-FBWs | 284 | 26 679 | 1.06 (0.94-1.20) | ref | - | ref | - |
| FBWs | 21 | 673 | 3.12 (1.93-4.77) | 3.15 (1.96-4.79) | <0.001 | 3.71 (2.30-5.67) | <0.001 |
| **Round 18-19** | 51 | 11 875 | 0.43 (0.32-0.56) |  |  |  |  |
| Non-FBWs | 48 | 11 583 | 0.41 (0.31-0.55) | ref | - | ref | - |
| FBWs | 3 | 292 | 1.03 (0.21-3.00) | 2.48 (0.60-6.75) | 0.127 | 2.99 (0.72-8.24) | 0.068 |

Abbreviations: FBW, female bar worker; HIV, human immunodeficiency virus; IR, incidence rate; CI, confidence interval; IRR, incidence rate ratio; PY, person-years

#### Supplementary Table 6. Incidence rates and incidence rate ratios of HIV seroconversion by self-reported history of ever engaged in bar work as a primary or secondary occupation among female participants in the Rakai Community Cohort Study, Uganda, 2011-2020 (N=9726)

| **Population** | **HIV seroconversion** | **Person-years** | **IR per 100 PY (95% CI)** | **IRR (95%CI)** | ***P*-value** | **Age-adjusted IRR (95%CI)** | ***P*-value** |
| --- | --- | --- | --- | --- | --- | --- | --- |
| **Overall** | 356 | 39 228 | 0.91 (0.82-1.01) |  |  |  |  |
| Non-FBWs | 317 | 37 714 | 0.84 (0.75-0.94) | ref | - | ref | - |
| FBWs | 39 | 1513 | 2.58 (1.83-3.52) | 3.23 (2.28-4.44) | <0.001 | 3.82 (2.69-5.29) | <0.001 |
| **Inland community** | 228 | 33 777 | 0.68 (0.59-0.77) |  |  |  |  |
| Non-FBWs | 206 | 32 736 | 0.63 (0.55-0.72) | ref | - | ref | - |
| FBWs | 22 | 1040 | 2.12 (1.33-3.20) | 3.57 (2.23-5.41) | <0.001 | 4.23 (2.63-6.48) | <0.001 |
| **Fishing community** | 128 | 5451 | 2.35 (1.96-2.79) |  |  |  |  |
| Non-FBWs | 111 | 4978 | 2.23 (1.83-2.69) | ref | - | ref | - |
| FBWs | 17 | 473 | 3.59 (2.09-5.75) | 1.61 (0.93-2.61) | 0.067 | 1.81 (1.05-2.94) | 0.023 |

Abbreviations: FBW, female bar worker; HIV, human immunodeficiency virus; IR, incidence rate; CI, confidence interval; IRR, incidence rate ratio; PY, person-years

#### Supplementary Table 7. Prevalence and prevalence ratios of HIV viral suppression by self-reported history of bar work as a primary or secondary occupation among HIV seropositive visits in the Rakai Community Cohort Study, Uganda, 2013-2020 (N=8763)

| **Population** | **Viral suppression, % (case/total)** | **PR (95%CI)** | ***P*-value** | **Age-adjusted PR (95%CI)** | ***P*-value** |
| --- | --- | --- | --- | --- | --- |
| **Overall** | 82.2 (7201/8763) |  |  |  |  |
| Non-FBWs | 82.2 (6610/8019) | ref | - | ref | - |
| FBWs | 81.9 (591/744) | 1.00 (0.96-1.04) | 0.849 | 0.99 (0.95-1.03) | 0.494 |
| **Inland community** | 82.8 (4320/5220) |  |  |  |  |
| Non-FBWs | 82.7 (4004/4841) | ref | - | ref | - |
| FBWs | 83.4 (316/379) | 1.02 (0.97-1.08) | 0.389 | 1.00 (0.95-1.05) | 0.997 |
| **Fishing community** | 81.3 (2881/3543) |  |  |  |  |
| Non-FBWs | 81.4 (2588/3178) | ref | - | ref | - |
| FBWs | 80.3 (293/365) | 0.99 (0.93-1.05) | 0.699 | 0.97 (0.92-1.03) | 0.332 |

Abbreviations: FBW, female bar worker; PR, prevalence ratio; CI, confidence interval.

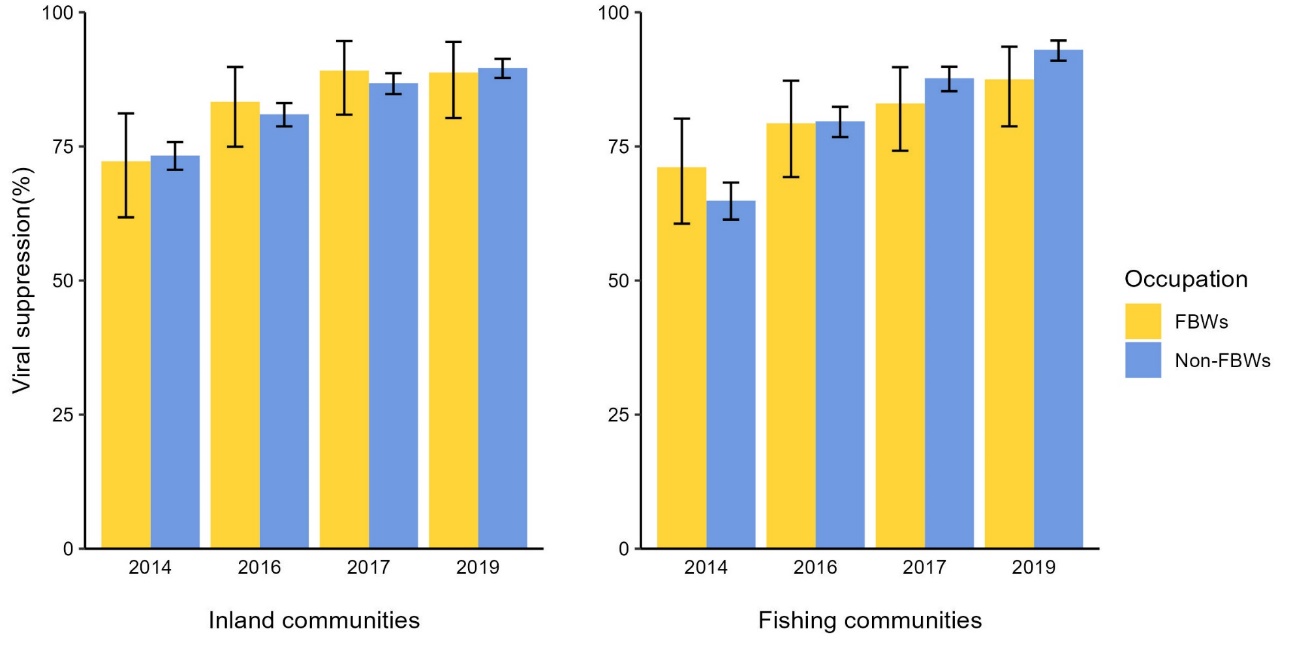

Abbreviation: FBW, female bar worker

#### Supplementary Figure 6. HIV viral suppression by self-reported history of bar work as a primary or secondary occupation and by survey round among HIV seropositive visits of Rakai Community Cohort Study, Uganda, 2013-2020 (N=8763)

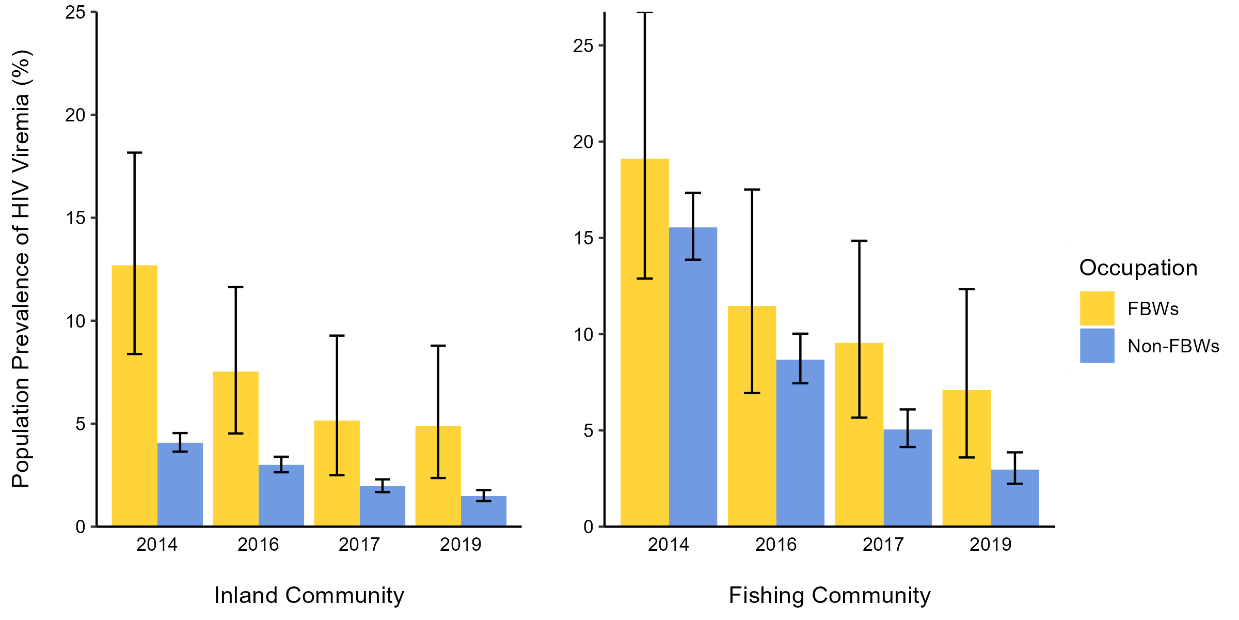

Abbreviation: Abbreviations: FBW, female bar worker; HIV, human immunodeficiency virus

#### Supplementary Figure 7. Population prevalence of HIV viremia by the self-reported history of bar work as a primary or secondary occupation and by community type at each survey round among female participants of Rakai Community Cohort Study, Uganda, 2013-2020 (N=20 250)

#### Supplementary Table 8. Awareness of HIV serostatus among women living with HIV of the survey round 19 of the Rakai Community Cohort Study, Uganda, 2018-2020 (N=2135)

| **Awareness of HIV serostatus** | **Non-FBWs** | **FBWs** | ***P-*value** |
| --- | --- | --- | --- |
|  | **N=1958** | **N=177** |  |
| No | 95 (4.9%) | 6 (3.4%) | 0.380 |
| Yes | 1863 (95.1%) | 171 (96.6%) |  |

Abbreviations: FBW, female bar worker; HIV, human immunodeficiency virus

#### Supplementary Table 9. PrEP awareness and use among HIV seronegative female participants of the survey round 19 of the Rakai Community Cohort Study, Uganda, 2018-2020 (N=7827)

| **Characteristics** | **Non-FBWs** | **FBWs** | ***P-*value** |
| --- | --- | --- | --- |
|  | **N=7648** | **N=179** |  |
| **PrEP awareness** |  |  | 0.032 |
| No | 2089 (27.3%) | 36 (20.1%) |  |
| Yes | 5559 (72.7%) | 143 (79.9%) |  |
| **PrEP use** |  |  | <0.001 |
| Never | 7436 (97.2%) | 155 (86.6%) |  |
| Previous | 103 (1.3%) | 19 (10.6%) |  |
| Current | 109 (1.4%) | 5 (2.8%) |  |

Abbreviations: FBW, female bar worker; HIV, human immunodeficiency virus; PrEP, Pre-exposure prophylaxis
